# Identification of *ATP2B4* regulatory element containing functional genetic variants associated with severe malaria

**DOI:** 10.1101/2021.12.03.21267245

**Authors:** Samia Nisar, Magali Torres, Alassane Thiam, Bruno Pouvelle, Florian Rosier, Frederic Gallardo, Oumar Ka, Babacar Mbengue, Rokhaya Ndiaye Diallo, Laura Brosseau, Salvatore Spicuglia, Alioune Dieye, Sandrine Marquet, Pascal Rihet

## Abstract

Genome-wide association studies (GWAS) for severe malaria have identified 30 genetic variants that are mostly located in non-coding regions, with only a few associations replicated in independent populations. In this study, we aimed at identifying potential causal genetic variants located in these loci and demonstrate their functional activity. We systematically investigated the regulatory effect of the SNPs in linkage disequilibrium with the tagSNPs associated with severe malaria in several populations. Annotating and prioritizing genetic variants led to the identification of a regulatory region containing 5 *ATP2B4* SNPs in linkage disequilibrium with the tagSNP rs10900585. We confirmed the association of rs10900585 and also found significant associations of severe malaria with our candidate SNPs (rs11240734, rs1541252, rs1541253, rs1541254, and rs1541255) in a Senegalese population. Then, we showed that this region had both promoter and enhancer activities and that both individual SNPs and the combination of SNPs had regulatory effects using luciferase reporter assays. Moreover, CRISPR/Cas9-mediated deletion of this region decreased *ATP2B4* transcript and protein levels and increased Ca^2+^ intracellular concentration in the K562 cell line. Taken together, our data show that severe malaria-associated genetic variants alter the activity of a promoter with enhancer function. We showed that this regulatory element controls the expression of *ATP2B4* that encodes a plasma membrane calcium-transporting ATPase 4 (PMCA4), which is the major calcium pump on red blood cells. Altering the activity of this regulatory element affects the risk of severe malaria probably through calcium concentration effect on parasitaemia.

## Introduction

Malaria caused by *Plasmodium falciparum* parasites is a major cause of morbidity and mortality in many developing countries predominantly in endemic areas of Sub-saharan Africa. The disease outcome is variable and may be subjective to a combination of various factors including host genetic factors, parasite virulence as well as environmental factors ^1,2^. *P. falciparum* infection causes various clinical phenotypes from asymptomatic parasitemia and uncomplicated malaria to severe malaria (SM) ^3^. There is a growing body of evidence that human genetic factors influence the outcome of infection since the discovery of the protective effect of the sickle cell mutant (rs334). Heritability studies reported that genetic factors explain from 20% to 25% of the variations in malaria phenotypes ^1,4,5^. Genetic linkage analyses conducted in Africa provided evidence of linkage of uncomplicated malaria with chromosome 6p21 ^6–8^, whereas parasitemia was genetically linked to chromosome 5q31-q33 ^5,6,9–12^. Moreover, several GWAS of SM have identified several malaria resistance loci ^13–18^. However, a limited number of loci and genetic variants were identified and replicated in GWAS: *ATP2B4* on chromosome 1, *FREM3-GYP A/B* on chromosome 4, *EPHA7* on chromosome 6, *ABO* on chromosome 9, and *HBB* on chromosome 11. rs334 that is a nonsense polymorphism is the main causal SNP within the *HBB* locus ^19^, and a copy number variant within *GYP A/B* locus was shown to confer protection against malaria in East Africa ^20^. The specific variant named DUP4 encodes hybrid glycophorin proteins that may alter the invasion of red blood cells by the parasite ^21,22^. Besides, it is thought that regulatory genetic variants alter SM resistance. In this way, Lessard et al (2017) evidenced that an enhancer containing rs10751450, rs10751451, and rs10751452 influences the expression of *ATP2B4*, which encodes for PMCA4 (Plasma membrane calcium transporting ATPase4), the major Calcium pump of red blood cells ^23^. Identifying causal variants from GWAS results remains challenging, although some important results have been obtained. In particular, it is thought that unknown non-coding variants may alter the expression of genes associated with malaria phenotypes.

Here we looked for regulatory SNPs in linkage disequilibrium (LD) with tagSNPs, for which association with SM has been replicated in independent populations. We prioritized and annotated the SNPs, and further investigated the best candidates for their functional activity. We provide evidence that several SNPs were associated with SM in a Senegalese population alter the activity of an Epromoter affecting the expression of *ATP2B4* and modulate intracellular calcium concentration.

## Materials and methods

### Study Subjects, blood samples, and Phenotypes

Malaria patients were recruited in two major Senegalese sites, the principal hospital of Dakar and the regional hospital of Tambacounda including 90 cerebral malaria cases and 27 severe noncerebral cases, as described ^24^. The control samples (*n*=79) were obtained from healthy volunteers living in Dakar. Written informed consent was obtained for each patient or their accompanying family members. The study was approved by the institutional research ethics committee of the Université Cheikh Anta Diop.

Venous blood samples and biological data including parasite density, hematology, and other characteristics were collected on the day of admission. The presence of *Plasmodium falciparum* was determined by at least two trained biologists by microscope examination of thin and thick smears before anti-malarial treatment. If the asexual parasites were observed, then the slide was considered as positive, and the number of parasitized erythrocytes was counted per µl of blood.

### Bioinformatic prioritization and functional annotation of genetic variants

Significant GWAS signals replicated at least in an independent population were identified and the corresponding tagSNPs (rs4951377, rs10900585, rs186873296, rs62418762, and rs8176719) were selected for further analysis. Haploreg v4.1, a tool including LD information from the 1000 Genome project was used to identify the SNPs in LD with these tagSNPs. This analysis of the African population and LD threshold r^2^>0.6 resulted in the selection of 125 SNPs. To prioritize the SNPs with putative functional significance, we performed bioinformatic analysis using the IW-Scoring annotation tool (https://www.snp-nexus.org/IW-Scoring/) ^25^ and Remap 2018 based on DNA-binding ChIP-seq experiments ^26^. We also checked whether these SNPs have been annotated as eQTLs (https://pubs.broadinstitute.org/mammals/haploreg/haploreg.php) or identified as regulatory SNPs using the massive parallel reporter assay named Sure-seq ^27^. Additional analysis has been performed to evaluate whether the candidate SNPs are located within a DNAse I hypersensitivity region, peaks of H3K4me1, H3K4me3, and H3K27ac marks in the K562 cell line and visualized them in the UCSC genome browser.

### DNA extraction, DNA amplification, and genotyping

Genomic DNA was extracted and amplified as previously described ^24^. In total, we genotyped 9 SNPs of *ATP2B4* including rs11240734, rs1541252, rs1541253, rs1541254 and rs1541255, the TaqSNP rs10900585 and the SNPs rs10751450, rs10751451 and rs10751452 previously described as regulatory variants ^23^. Genotyping was performed using either the Sanger sequencing or the TaqMan SNP genotyping assays (Applied Biosystems, ThermoFisher). All the primer pairs for sequencing were designed using Primer 3 software. PCR amplification of DNA fragments containing rs11240734, rs1541252, rs1541253, rs1541254 and rs1541255 was performed with forward (5’-TCAGGCCTAGCTATCAGTTCAG-3’) and reverse (5’-CGAGTAGCCGTCCGAAGTC-3’) primers, respectively. For the tagSNP rs10900585, the forward and the reverse primers were (5’-GGGATGAGGAGGCTTACAGG-3’) and (5’-GAGGTTGAGGTGAGCGGATC-3’), respectively.

PCR amplification was carried out in 50µl reaction volume with 12.5 ng of genomic DNA, GoTaq G2 2X ready to use Master Mix (Promega, cat#M7433), and 10 µM of each primer. For rs10900585, annealing temperature was at 65°C while for rs1541252, annealing was at 60°C. The PCR products were submitted to electrophoresis on 2% agarose gel to verify the product size and then purified with a PCR Purification kit (Qiagen, Hilden, Germany) following the manufacturer’s protocol. Five µl of purified PCR products were sequenced using the Sanger method (GATC biotech, Germany, or Eurofins Genomics, Germany).

For rs10751450, rs10751451 and rs10751452, we used TaqMan genotyping assays C_31796478_10, C_31796479_10 and C_31796480_10, respectively. PCR amplification was performed with the QuantStudio 6 Flex Real-Time PCR Systems (Applied Biosystem, ThermoFisher Scientific) in a final volume of 5 µl containing 2.1 µl of Master Mix (Applied Biosystem, ThermoFisher Scientific), 0.06 µl of TaqMan probe and 1.84 µl H_2_O and 1 µl of DNA (10-15 ng/µl).

### Luciferase Gene Reporter Assay and site-directed mutagenesis: Promoter activity

A 601 bp DNA fragment upstream the *ATP2B4* translation start site (chromosome 1: 203682549-203683049 according to the hg38 assembly) was cloned into the MlulI-XhoI sites of the pGL3-basic vector (Promega, Madison, WI, USA, cat# E1751), which contained the firefly luciferase coding sequence (GeneCust Custom Services, Luxembourg). We obtained two different pGL3 constructs containing either minor or major alleles of the 5 SNPs (rs11240734, rs1541252, rs1541253, rs1541254, and rs1541255). From the minor allele construct, we generated additional constructs by replacing the minor allele with the major allele of each SNP individually. The site-directed mutagenesis was performed using the Q5 Site-Directed Mutagenesis Kit (New England Biolabs, Ipswich, MA, USA, cat#E0554S) and primers designed by NEBaseChanger tool provided by the supplier (detail sequences provided in (Table S1). K562 cells (ATTC CLL-243) were grown in Gibco RPMI 1640 medium (Thermo Fisher Scientific Waltham, MA, USA) supplemented with 10% of FBS (fetal bovine serum). K562 transfection was performed with the Neon^TM^ Transfection system (Invitrogen) according to the manufacturer’s instructions. For each out of 9 tests performed, 10^6^ cells were co-transfected with 1µg of control vectors 1) negative control vector (empty pGL3-basic vector (cat# E1751)) or 2) positive control vector (pGL3-promoter vector (cat# E1761)) or with 1 µg of the construct to be tested (a pGL3-basic vector containing either the minor alleles CTTCG or the major alleles TCCGA or one of these combinations TTTCG, CCTCG, CTCCG, CTTGG, and CTTCA) and 200 ng of pRL-SV40 (a plasmid encoding renilla luciferase from Promega (cat# E2231)), which was used as a transfection efficiency control. Transfected cells were maintained at 37°C in 5% CO_2_ for 24 hours. Values of Firefly and renilla luciferase were obtained by analyzing 20 µl of cell lysate according to the standard instructions provided in the Dual-Luciferase kit (Promega cat# E1910) in a TriStar LB 941 Multimode Microplate Reader (Berthold Technologies, Thermo Fisher Scientific, Waltham, MA, USA). Firefly luciferase activity of each sample was normalized to renilla luciferase and expressed as the fold change of the empty vector control.

### Enhancer activity

A 780 bp fragment of *ATP2B4* promoter (chromosome1: 203626081-203626860 (hg38 assembly)) was cloned into pGL3-Enhancer vector (Promega, cat#E1771, Promega) between MluI-Xho I sites and the 601 bp fragment of *ATP2B4* enhancer (chromosome 1: 203682549-203683049 according to the hg38 assembly) containing respective minor and major alleles of the 5 SNPs was cloned into BamHI-SalI sites. Luciferase assays were performed in K562 cells as described above.

### CRISPR-Cas9 genome editing in K562 cells

Using CRISPR design tool provided by IDT, we identified guide RNAs to target *ATP2B4* specific region. Chemically synthesized oligoribonucleotides were manufactured by IDT: the crRNAs (35 mer with specific part to DNA target sequence) and the universal tracrRNAs (67 mer). A two-part system where synthetic crRNA (5 µl of 100 µM) and tracrRNA (5 µl of 100 µM) were annealed to form an active gRNA complex. Cas9 RNP complexes were assembled in vitro by incubation of 3.4 µl Cas9 protein (62 mM) with 4.8 µl active gRNA complex (cr:tracrRNA) and 1.8 µl of PBS. To generate the genomic deletion two different RNP complexes were simultaneously electroporated with the Neon transfection system into 5.10^5^ K562 cells. The small deletion of 506 bp (containing the 5 SNPs) was obtained by using 2 µl gRNA1 and 2 µl of gRNA2 (details of sequences in (Table S2). The large deletion of 1262 bp (containing the 8 SNPs) was performed by using gRNA2 and gRNA3 (Table S2). Those bulk cultures transfected with tandem gRNA were plated clonally in 96-well plates at limiting dilution less than 0.5 to avoid mixed clones. After approximately 14 days of clonal expansion, amplification of DNA was performed directly from the clones by using the 2X Phire Tissue Direct PCR Master Mix containing Phire Hot Start II DNA Polymerase (Thermo Fisher Scientific), with 20µl of dilution buffer and 0.5µl of DNA Release additive, which allowed to improve the release of DNA from the cells. Clones were screened for deletion by PCR using primers F1 and R1 for the small deletion detection that would produce a specific short fragment of 335 bp in the presence of deletion and a long fragment of 841 bp in the absence of deletion (Table S2). Primers F2 and R2 were used for large deletion detection that would produce amplicons of 355 bp in presence of deletion and 1617 bp in the absence of deletion (Table S2). PCR was performed using 10 µl of 2x Master Mix and 10 µM of each primer, using annealing at 60°C. After electrophoresis on 1% agarose gel, we selected homozygotes deleted clones showing only amplicons of 335 bp or 355 bp for the small and the large deletions, respectively. After sequencing using the Sanger method only the clones with the appropriate sequence with no additional edits were kept for gene expression analysis.

### Reverse transcription-quantitative PCR

Total RNA was extracted for each selected clone and wild-type K562 cells using the RNeasy mini kit (Qiagen, Hilden, Germany). One µg of RNA per clone was converted into cDNA using Superscript VILO Master Mix (Invitrogen, ThermoFisher Scientific). Three independent RNA/cDNA preparations were performed for each clone. Real-time quantitative PCR (RT-qPCR) was subsequently performed using SYBR Select Master Mix (ThermoFisher Scientific) on a QuantStudio 6 Flex instrument. Primers were designed using Primer 3 software to span exon 1 of *ATP2B4* (F3 and R3) to specifically quantify the long transcripts (ENST00000357681, ENST00000367218) or to span exon 19 and exon 20 (F4 and R4) to quantify all the *ATP2B4* transcripts (ENST00000357681, ENST00000367218, ENST00000341360, ENST00000458092, and ENST00000356729) (Table S3). PCR efficiency was validated by performing serial dilution analysis. Gene expression was normalized to that of actin and the relative expression was calculated by the ΔCT method. The data reported correspond to the mean of triplicates from three independent experiments per clone and expressed as fold change relative to wild-type cells. Three independently generated clones were analyzed for gene expression quantification of *ATP2B4*.

### Calcium measurement by flow cytometry in K562 clones and wild-type cells

The intracellular quantity of calcium was measured in K562 cells and ΔATP2B4_2 and ΔATP2B4_3 clones, by flow cytometry, using the Fluo-4, AM kit (Molecular Probes/ThermoFisher Scientific), following manufacturer’s instructions. Briefly, samples of 10^6^ cells were harvested for each cell type. The cells were pelleted and resuspended in 1 ml of culture medium without serum. One ml of either Fluo-4-AM (labeled cells) or kit buffer (control cells) was added to the cells, before 1 hour incubation at 37°C. The cells were then directly observed on an LSRFortessa X-20 cytometer (BD Biosciences). Cells were gated on forward/side-light scatter and the fluorescence intensity of 20,000 cells was measured at the FITC excitation/emission wavelength, for each sample.

### Detection of PMCA4 by flow cytometry in K562 clones and wild-type cells

The presence of PMCA4 in K562 cells and clones ΔATP2B4_2 and ΔATP2B4_3 was detected by flow cytometry, using the specific antibody JA9 (ABCAM, ab2783). The labeling was realized according to a slight modification of the manufacturer’s protocol. For each cell type, 2×10^6^ cells were harvested, pelleted, and fixed in 2 ml of 80% methanol (5 min). After centrifugation, the cells were incubated in 2 ml of 1xPBS / 1% BSA, for 30 min at room temperature, to block non-specific interactions. Half of each cell type was then incubated with the JA9 antibody (2 µg/1×10^6^ cells) for 30 min at room temperature. The other half was used for isotypic control (ab170190, 2 µg/1×10^6^ cells). After centrifugation, the cells were incubated with the secondary antibody (goat anti-mouse IgG1 APC, ab 130786) at 1/200 dilution in 1xPBS / 1% BSA, for 30 min at room temperature. Finally, cells were washed twice in 1xPBS and observed on an LSRFortessa X-20 cytometer (BD Biosciences). Cells were gated on forward/side-light scatter and the fluorescence intensity of 20 000 cells was measured at the APC excitation/emission wavelength, for each sample.

### Statistical analysis

A Chi2 test was used to determine whether the genotype distribution in healthy Senegalese subjects conformed to Hardy-Weinberg equilibrium. Chi2 test and logistic regression analysis were carried out with SPSS (statistical software version 20) to assess the association between the SNPs and SM in Senegalese subjects. Differences were considered significant if the *P*-value obtained in a two-tailed test was < 0.05. The odds ratio (OR), 95% confidence intervals (CIs), and the influence of covariates on the phenotype were evaluated by logistic regression analysis. A meta-analysis of the genetic association was performed using MetaGenyo ^28^. Statistical differences between luciferase constructs or between clones were performed using Student’s *t*-tests. Mixed models were also used to take into account the triplicates performed for each independent experiment. *P* values <0.05 were considered statistically significant. Plot generation was performed using GraphPad Prism. Statistical analysis to evaluate the difference in the calcium concentration between deleted and wild-type clones was based on the non-parametric Wilcoxon test using SPSS, and on a meta-analysis approach using Open Meta-Analyst software ^29^. All the tests used were two-sided tests.

### Results Prioritization and annotation of putative regulatory SNPs in linkage disequilibrium with tagSNPs associated with severe malaria

We focused our analysis on the SNPs, for which significant GWAS signals have been replicated: rs4951377 and rs10900585 at the *ATP2B4* locus, rs186873296 at the *FREM3-GYP A/B* locus, rs62418762 at the *EPHA7* locus, and rs8176719 at the *ABO* locus. We excluded the coding variation rs334, which captured the GWAS signal at the *HBB* locus. We identified 126 SNPs in LD with the malaria-associated SNPs, based on an r^2^ higher than 0.6. We excluded rs181620317 that is a missense variant within the *FREM3* coding sequence, and further prioritized the SNPs for their potential regulatory effect using the IW-scoring ^25^. Moreover, we investigated the ability of sequences containing the SNPs to bind transcription factors using the Remap tool ^26^, and whether the 125 SNPs have been annotated as eQTLs or identified as regulatory SNPs ^27^. (Table S4). As shown in Figure 1A, the number of peaks identified by ChIP-seq negatively correlated with the rank of the corresponding SNPs (rho=-0.652, P=2.17 10^−11^). The highest numbers of peaks were observed for rs1541252, rs1541253, rs1541254, and rs1541255 (*n*>90), which were ranked second, 10^th^, 6^th^, and 7^th^ based on the IW-score, respectively (Fig. 1B). The best SNP according to the IW-scoring method was rs11240734, for which 54 peaks of ChIP-seq were registered in the Remap catalog (Fig. 1B). Strikingly, those 5 SNPs are close to each other on chromosome 1 in the *ATP2B4* region. These SNPs are located within a DNAse I hypersensitivity region, and peaks of H3K4me1, H3K4me3, and H3K27ac marks in the K562 cell line, as shown in Figure 2. Similar epigenomic marks have been found at these positions in erythroblasts (Fig. S1). rs11240734 and rs1541252 have been annotated as eQTLs, according to Haploreg, whereas rs1541255 was found to have a regulatory effect, according to the Sure-seq method ^27^.

**Figure 1.**
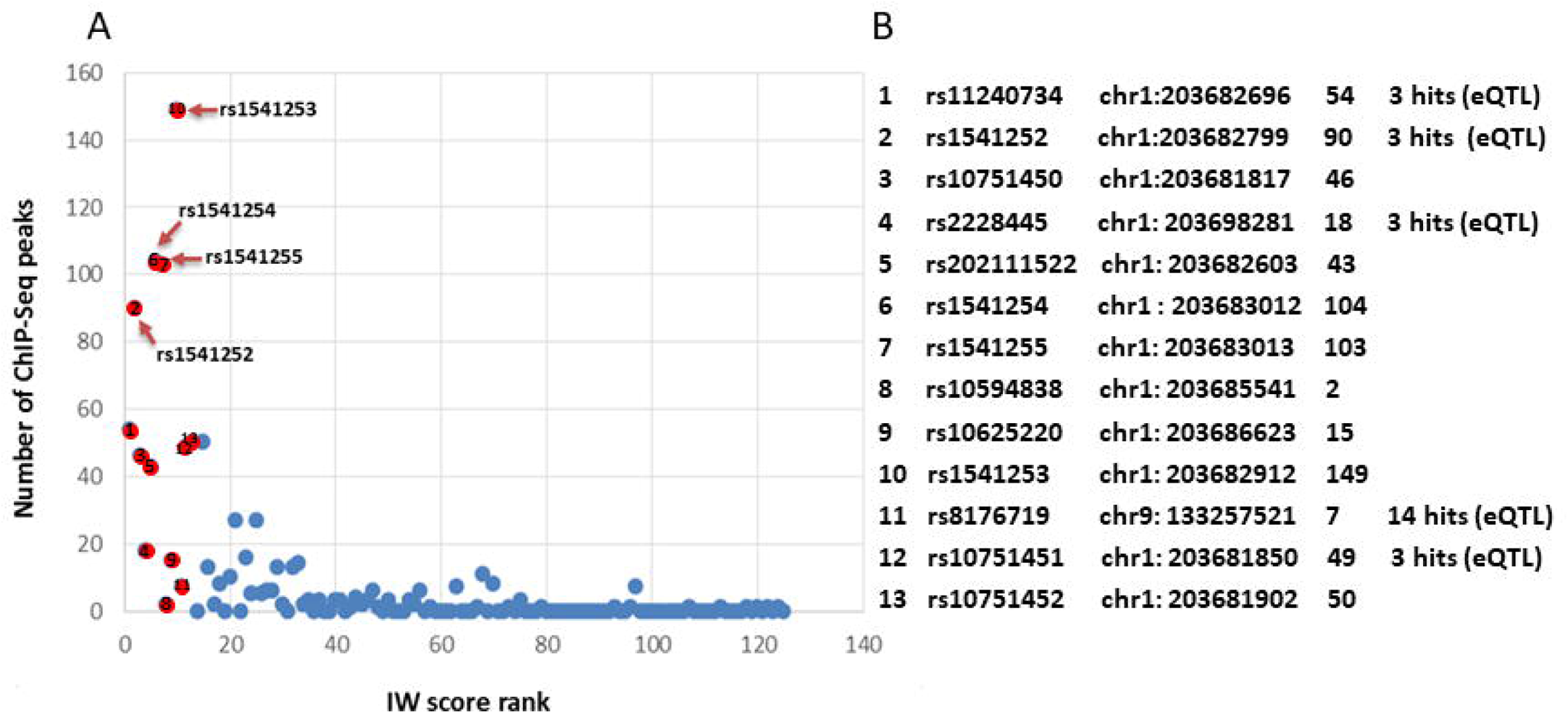
Plot showing the integrated results for the prioritization of best candidate SNPs. **(A)** The prioritization was based on IW scoring analysis on the X-axis and no. of ChIP-seq peaks on the Y-axis. The top 13 SNPs according to the IW-score are shown in red and numbered according to their rank. The 4 SNPs rs1541252, rs1541253, rs1541254, and rs1541255 showing the highest number of ChIP-seq peaks (*n*>90) are shown. **(B)** Information on SNPs rsID, genomic coordinate (hg38), number of chip-seq peaks and eQTL hits are indicated for the Top13 SNPs.

**Figure 2.**
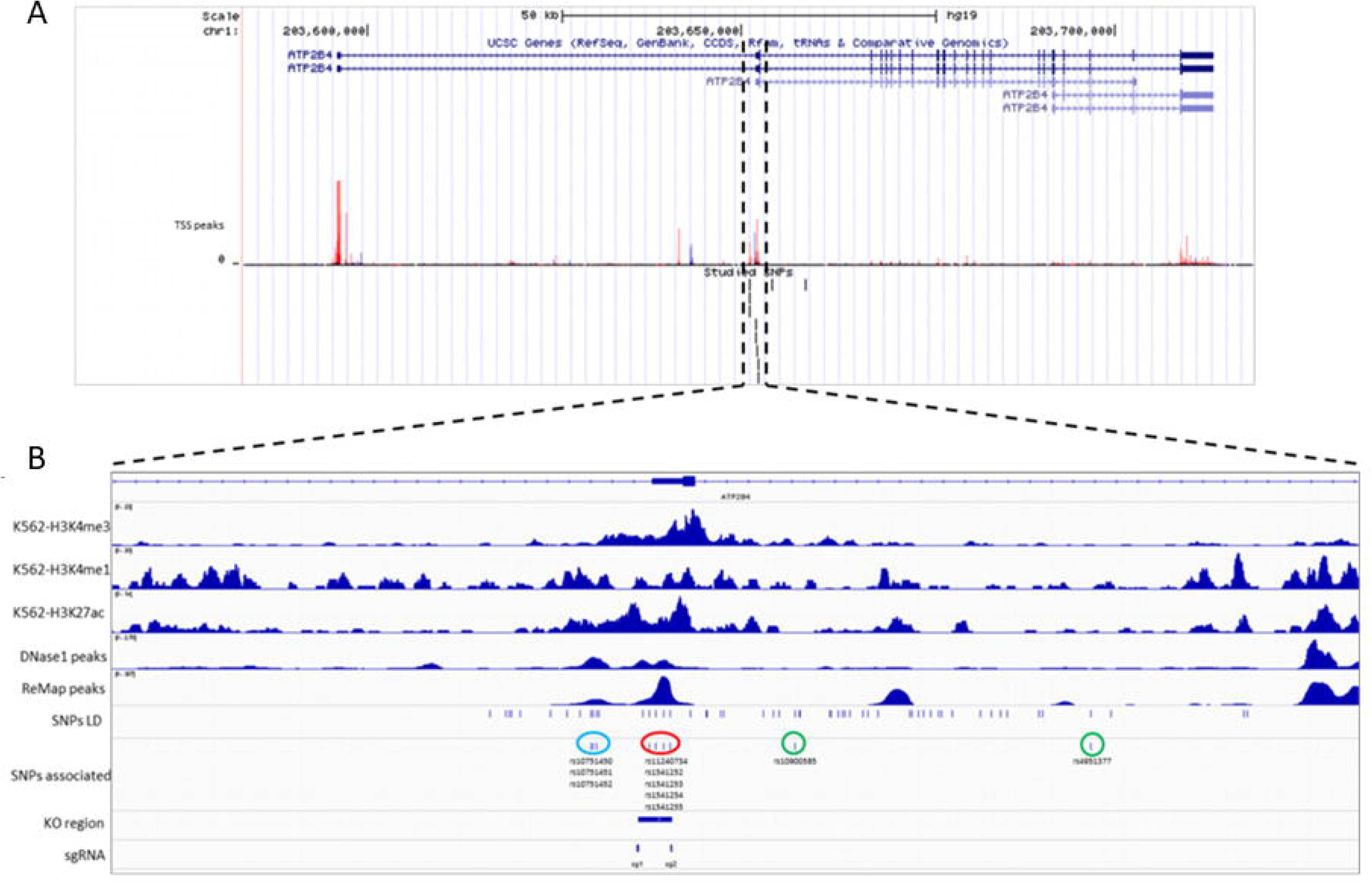
Visual representation of the *ATP2B4* locus and the epigenomic marks of the region containing the 5 *ATP2B4* candidate variants. **(A)** Transcripts, TSS peaks, and studied SNPs are shown. The two long transcripts (*ATP2B4*-203 and *ATP2B4*-204), and the short transcripts are shown. The main TSS peaks are visualized. **(B)** The candidate SNPs (rs11240734, rs1541252, rs1541253, rs1541254, and rs1541255; encircled red) are on chromosome 1, located within a DNaseI hypersensitivity region and peaks of H3K4me3, H3K4me1, and H3K27ac histone marks. The 2 tagSNPs (rs10900585 and rs4951377; encircled green) are located neither in peaks of ChIP-seq in the ReMap catalog nor in other epigenomic marks. The three additional regulatory variants (rs10751450, rs10751451 and rs10751452; encircled blue) previously identified as functional SNPs are shown.

Interestingly, the TaqSNPs rs4951377 and rs10900585 that are in LD with rs11240734, rs1541252, rs1541253, rs1541254, and rs1541255 had a low IW-score and were ranked at positions 51 and 60, respectively (Table S4). Furthermore, there were neither peaks of ChIP-seq in the Remap catalog nor peaks of H3K4me1, H3K4me3, and H3K27ac marks in the K562 cell line for both SNPs (Fig. 2), suggesting that they have no functional role. In contrast, rs10751450 ranked third with 46 peaks of Chip-Seq is located within a DNAse I hypersensitivity region, and peaks of H3K4me1 and H3K27ac marks in the K562 cell line (Fig. 2), and is located within a regulatory region ^23^. Similar epigenomic profiles were observed in erythroblasts (Fig.S1). The SNPs rs10751451 and rs10751452 close to the rs10751450 ^23^ were ranked 12^th^ and 13^th^, and rs10751451 was annotated as an eQTL according to Haploreg (Fig. 1B). Figure 2 shows the position of the region containing rs10751450, rs10751451, and rs10751452 and that containing rs11240734, rs1541252, rs1541253, rs1541254, and rs1541255. Overall, SNP prioritization and annotation allowed us to identify 5 new SNPs with potential regulatory effect, which may drive the observed association of their tagSNPs with malaria susceptibility.

### Association between the *ATP2B4* tagSNP rs10900585 and severe malaria in a Senegalese cohort

One hundred and ninety-six individuals from the Senegalese cohort, were successfully genotyped by sequencing (Table 1). The frequencies of observed genotypes conformed to the Hardy-Weinberg Equilibrium (*P*=0.52) (Table S5) and the allelic frequencies for rs10900585-T and G were found to be 0.59 and 0.41, respectively. We detected a significant association (*P*= 0.029; OR= 1.94) of *ATP2B4* rs10900585 with SM (Table 1) without the effect of any covariate. The TT genotype that was more frequent among cases (48.7%) than controls (32.9%) was identified as the risk genotype. Nevertheless, there was no significant association when taking into account age as a covariate (*P*= 0.055) (Table S6).

**Table 1:**
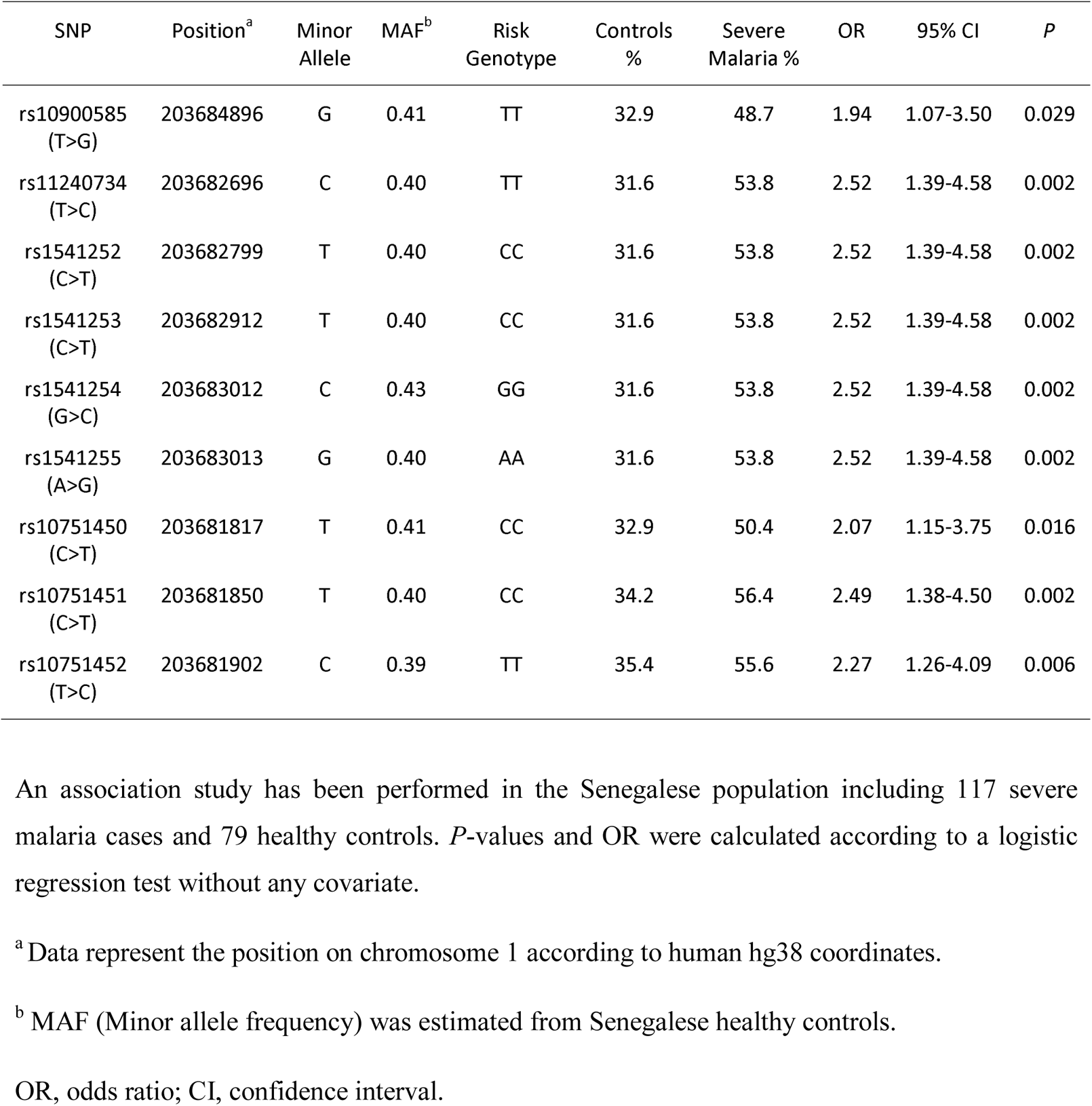
Association of *ATP2B4* SNPs with Severe Malaria in the Senegalese population-based study

| SNP | Position<sup>a</sup> | Minor Allele | MAF<sup>b</sup> | Risk Genotype | Controls % | Severe Malaria % | OR | 95% CI | *P* |
| --- | --- | --- | --- | --- | --- | --- | --- | --- | --- |
| rs10900585 (T>G) | 203684896 | G | 0.41 | TT | 32.9 | 48.7 | 1.94 | 1.07-3.50 | 0.029 |
| rs11240734 (T>C) | 203682696 | C | 0.40 | TT | 31.6 | 53.8 | 2.52 | 1.39-4.58 | 0.002 |
| rs1541252 (C>T) | 203682799 | T | 0.40 | CC | 31.6 | 53.8 | 2.52 | 1.39-4.58 | 0.002 |
| rs1541253 (C>T) | 203682912 | T | 0.40 | CC | 31.6 | 53.8 | 2.52 | 1.39-4.58 | 0.002 |
| rs1541254 (G>C) | 203683012 | C | 0.43 | GG | 31.6 | 53.8 | 2.52 | 1.39-4.58 | 0.002 |
| rs1541255 (A>G) | 203683013 | G | 0.40 | AA | 31.6 | 53.8 | 2.52 | 1.39-4.58 | 0.002 |
| rs10751450 (C>T) | 203681817 | T | 0.41 | CC | 32.9 | 50.4 | 2.07 | 1.15-3.75 | 0.016 |
| rs10751451 (C>T) | 203681850 | T | 0.40 | CC | 34.2 | 56.4 | 2.49 | 1.38-4.50 | 0.002 |
| rs10751452 (T>C) | 203681902 | C | 0.39 | TT | 35.4 | 55.6 | 2.27 | 1.26-4.09 | 0.006 |

An association study has been performed in the Senegalese population including 117 severe malaria cases and 79 healthy controls. *P*-values and OR were calculated according to a logistic regression test without any covariate.
<sup>a</sup> Data represent the position on chromosome 1 according to human hg38 coordinates.
<sup>b</sup> MAF (Minor allele frequency) was estimated from Senegalese healthy controls.
OR, odds ratio; CI, confidence interval.

We added 13 independent populations to our study population for performing a meta-analysis including 12,794 SM cases and 19,898 controls (Table S7) ^18,30,31^. There was no deviation from Hardy Weinberg equilibrium (*P*>0.05) but a significant heterogeneity among the studies (I^2^=66%; tau^2^=0.0174; P=0.0003). Thus, the random effect model was used for calculating the combined Odd Ratio and 95% confidence interval (CI). There was an association of rs10900585 with SM under the genetic dominant model (OR=1.12; 95% CI= 1.02-1.23) (Fig. 3A).

**Figure 3.**
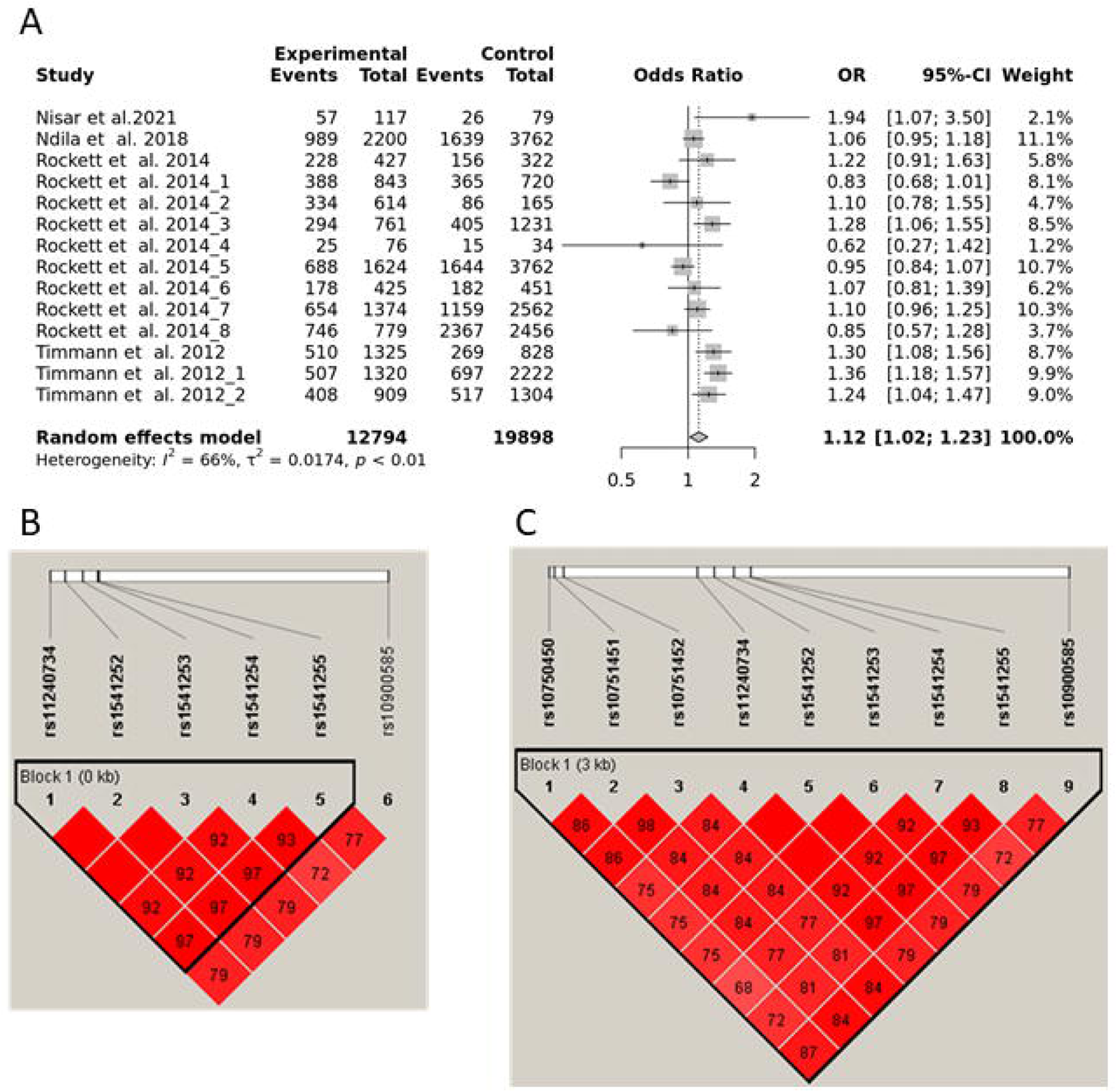
Meta-analysis and linkage disequilibrium. **(A)** Forest plot showing meta-analysis for the association between rs10900585 and SM. **(B, C)** Linkage disequilibrium (LD) (r-squared) plot for the *ATP2B4* variants in the Senegalese population. **(B)** LD between the candidate 5 SNPs and the tagSNP rs10900585 **(C)** LD between the 8 candidate SNPs and the tagSNP rs10900585 displaying r2 from 0.72-1.

### Association of rs11240734, rs1541252, rs1541253, rs1541254, and rs1541255 with severe malaria in Senegalese population

These 5 SNPs newly identified as potential regulatory variants were in a 600 bp DNA fragment that we sequenced to determine the genotypes of subjects. None of the polymorphisms deviated from the Hardy-Weinberg equilibrium (Table S5). We compared subjects carrying the homozygous genotype for the major alleles with those carrying either the heterozygous genotype or the homozygous genotype for the minor alleles. All these SNPs were significantly associated with the disease (*P* value= 0.002), with an estimated OR of 2.52 (1.39-4.58) (Table 1), indicating a stronger association compared to the tagSNP. Among the SM individuals, 53,8% carried the rs11240734TT, the rs1541252CC, the rs1541253CC, the rs1541254GG, and the rs1541255AA genotypes whereas only 31,6% of the control individuals carried these genotypes. The association of the 5 SNPs with SM remained significant (*P*=0.006) after taking into account age as a covariate (Table S6). In addition, we found a strong LD between all the 5 SNPs as well as with the tagSNP with an r^2^ varying from 0.72 to 1 (Fig. 3B).

### Association of rs10751450, rs10751451, and rs10751452 with severe malaria in Senegalese population

Although suggested as regulatory polymorphisms ^23^, the SNPs rs10751450, rs10751451, and rs10751452 have never been genotyped and tested for their association with malaria. We then tested whether these 3 SNPs were associated with SM. The frequencies of observed genotypes conformed to the Hardy-Weinberg Equilibrium (Table S5). The analysis indicated a significant association between malaria disease and these SNPs, rs10751450 (*P*=0.016; OR 2.07), rs10751451 (*P*=0.002; OR 2.49) and rs10751452 (*P*=0.006; OR 2.27) (Table 1) that remained significant when taking into account age as a covariate (Table S6). Noticeably, in our study population, these SNPs were found to be in strong LD between them as well as with our 5 new candidate SNPs (Fig. 3C).

### Haplotype analysis of the *ATP2B4* SNPs

Haplotype analysis, combining rs11240734, rs1541252, rs1541253, rs1541254, and rs1541255 revealed 5 haplotypes showing various frequencies, 0.65 for haplotype (1) with the major alleles (rs11210734T-rs1541252C-rs1541253C-rs1541254G-rs1541255A), 0.33 for haplotype (2) with the minor alleles (rs11210734C-rs1541252T-rs1541253T-rs1541254C-rs1541255G), 0.015 for the haplotype (3) (rs11210734T-rs1541252C-rs1541253C-rs1541254C-rs1541255A), 0.002 for haplotype (4) (rs11210734C-rs1541252T-rs1541253T-rs1541254C-rs1541255) and 0.002 for haplotype (5) (rs11210734C-rs1541252T-rs1541253T-rs1541254G-rs1541255A). We first investigated the relationship between SM and the two most frequent haplotypes (1 and 2). The haplotype (1) was found to be associated with an increased risk of developing SM (*P*=0.005). Among SM subjects, 55.8 % were homozygous for the susceptibility haplotype (1) and 34.5 % were heterozygous whereas only 31.6 % of the control subjects carried this haplotype at homozygote state. Individuals homozygous for haplotype (1) with major alleles have a higher risk of developing SM (*P*=0.002, OR=2.52) compared to heterozygous and homozygous haplotypes for minor alleles (Table 2). This association remained significant when including age as a covariate in the statistical model (*P*= 0.006) (Table 2). We further performed the haplotype analysis by including rs10751450, rs10751451, rs10751452, rs11240734, rs1541252, rs1541253, rs1541254 and rs1541255. Among SM individuals, 47.8% were homozygous and 27.4% were heterozygous for the haplotype (CCTTCCGA) while only 26.6 % of the control subjects carried this haplotype in the homozygous state. We found that haplotype with major alleles (CCTTCCGA) at the homozygous state was associated with an increased risk of SM (*P*=0.003, OR=2.67), even when taking age as a covariate (*P*=0.005) (Table 2).

**Table 2:** Haplotype association analysis for the 5 and 8 SNPs in the Senegalese population

| Major haplotype | Minor haplotype | Risk haplotype | *P*-value | SMvsCtrl |  |
| --- | --- | --- | --- | --- | --- |
|  |  |  |  | OR | 95% CI |
| TCCGA | CTTCG | TCCGA/TCCGA | 0.002<sup>a</sup> | 2.52<sup>a</sup> | 1.39-4.58<sup>a</sup> |
|  |  |  | 0.006<sup>b</sup> | 2.38<sup>b</sup> | 1.29-4.40<sup>b</sup> |
| CCTTCCGA | TTCCTTCG | CCTTCCGA/CCTTCCGA | 0.003<sup>a</sup> | 2.67<sup>a</sup> | 1.41-5.04<sup>a</sup> |
|  |  |  | 0.005<sup>b</sup> | 2.53<sup>b</sup> | 1.32-4.86<sup>b</sup> |

Haplotype analysis has been performed on the two most frequent haplotypes including severe malaria cases and healthy controls. *P*-values and OR were calculated according to logistic regression.
<sup>a</sup> The statistical model comparing individuals homozygous for major allele haplotype with individual heterozygous or homozygous for minor haplotype without any covariate.
<sup>b</sup> The statistical model by taking into account age as a covariate.
OR, odds ratio; CI, confidence interval.

### Lower promoter activity and higher enhancer activity are associated with the risk haplotype of severe malaria

Luciferase reporter assays were performed in K562 cell lines to evaluate the regulatory effect of the 5 SNPs on *ATP2B4* expression. Interestingly, this regulatory region showed a strong promoter activity compared to the SV40 promoter for both the risk haplotype with major alleles (*P*<1.10^−4^, 10.9-fold increase) and the non-risk haplotype with minor allele (*P*<1.10^−4^, 17.7-fold increase) (Fig. 4A). Moreover, the risk haplotype was shown to have reduced transcriptional activity than the non-risk haplotype (*P*<1.10^−4^). Hence, our results revealed differential allelic promoter activity in the K562 cell line showing that the non-risk haplotype exhibited 1.6-fold increased transcriptional activity compared to the risk haplotype. In addition, we observed that each SNP has a specific impact on expression and their combination reflects the combined effect observed. These results indicate that these 5 SNPs may be functional variants and that we must consider that it is their combined effect, which is probably involved in the susceptibility to severe forms as suggested also by the haplotypes naturally present in individuals living in endemic areas.

**Figure 4.**
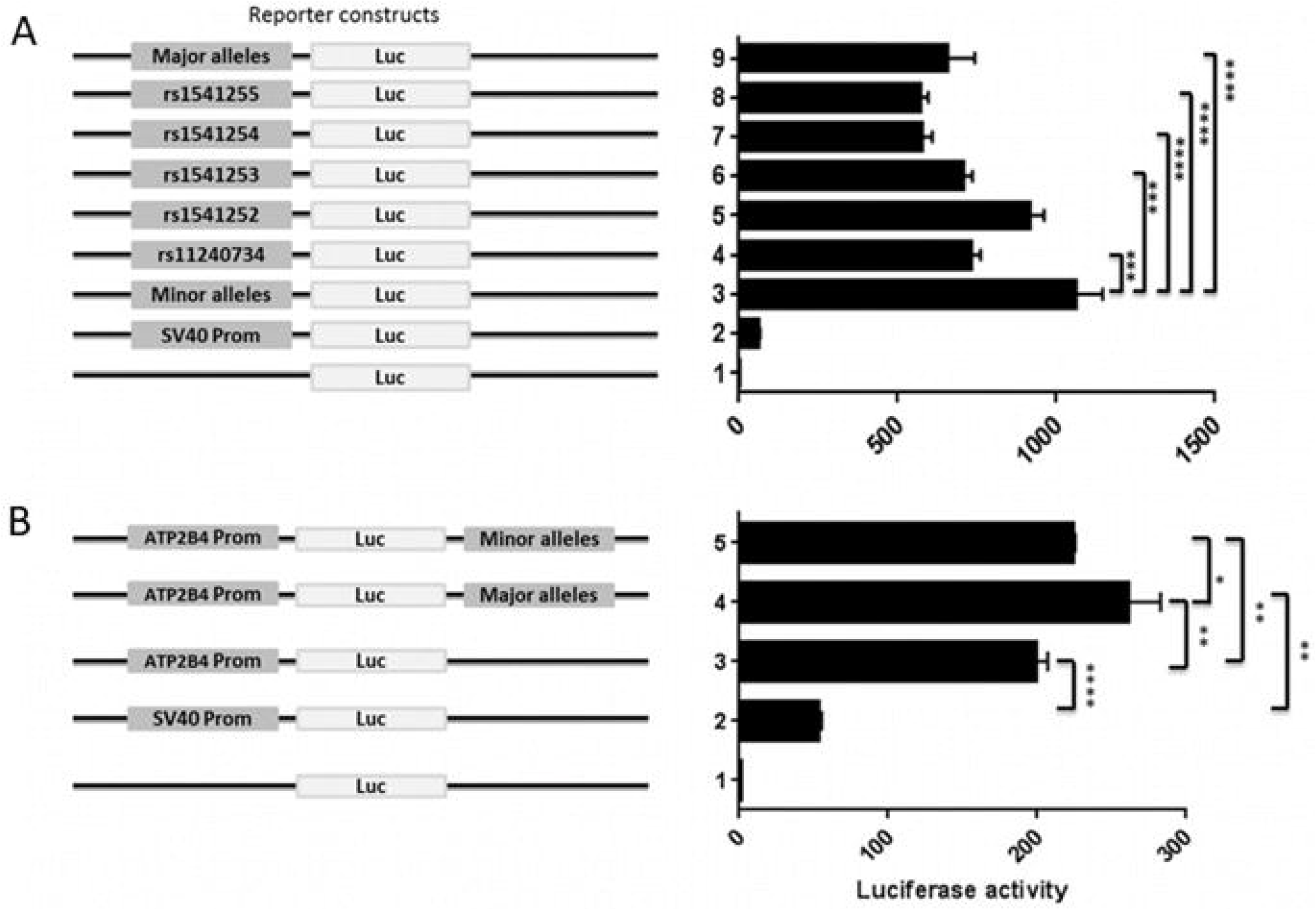
Luciferase reporter assay accessing the promoter and enhancer activity of *ATP2B4* variants in K562 cells. **(A)** Promoter activity of the DNA region containing *ATP2B4* Major and Minor haplotypes as well as the individual polymorphism. **(B)** Enhancer activity of *ATP2B4* SNPs with Major and Minor haplotypes. SV40 Promoter and Basic vector were used as positive and negative controls respectively. Data presented as the mean ± standard deviation of three independent experiments. \*\*\*\**P*<0.0001, ***P<0.001, \*\**P*<0.01, \**P*<0.05.

To functionally confirm that this genomic region may have enhancer function, we performed luciferase promoter assays by cloning it downstream the luciferase reporter gene derived by a promoter region of *ATP2B4* of 780 bp (Fig. 4B). We first validated the promoter activity of this 780 bp region compared to the SV40 promoter (*P*<1.10^−4^, 3.8-fold increase). We further confirmed the enhancer activity of the 600 bp region (*P*<0.01, 1.3-fold increase for the risk haplotype as compared to *ATP2B4* promoter). Moreover, we showed that the region containing the major alleles exhibited an enhancer activity that was significantly higher than that of the region containing the minor alleles (*P*=0.01). Also, there was a 1.2-fold increase in activity of the major allele haplotype relative to the minor allele haplotype. These data confirm that this regulatory region exhibits allele-dependent enhancer activity in the K562 cell line. Altogether, our results suggested that this regulatory region is a promoter with enhancer function, also named Epromoter ^32,33^. Furthermore, they suggest that genetic variants within this regulatory region orient the function of the region towards a promoter activity or towards an enhancer activity.

### Genome editing confirmed the regulatory activity of the region containing the SNPs

To analyze the function of the Epromoter element, we deleted a DNA region of 506-bp containing the malaria-associated SNPs (rs11240734, rs1541252, rs1541253, rs1541254, and rs1541255) in the K562 cell line, using the CRISPR/Cas9 mediated genome editing (Fig. 5A). We selected 3 deleted clones with the expected sequence and no additional edits. Expression analysis for total *ATP2B4* including long and short transcripts (Fig. 2) showed a 3.3-fold decrease in gene expression in deleted clones as compared to WT cells (P<1.10^−4^) (Fig. 5B). We also estimated the expression of the two long transcripts shown in Figure 2 (*ATP2B4*-203 and *ATP2B4*-204) of *ATP2B4* (encoding for PMCA4a and PMCA4b) and we showed a 2.8-fold decrease in deleted clones compared to WT cells (*P*<1.10^−4^) (Fig. 5B). These results support the hypothesis that this region has an enhancer function.

**Figure 5.**
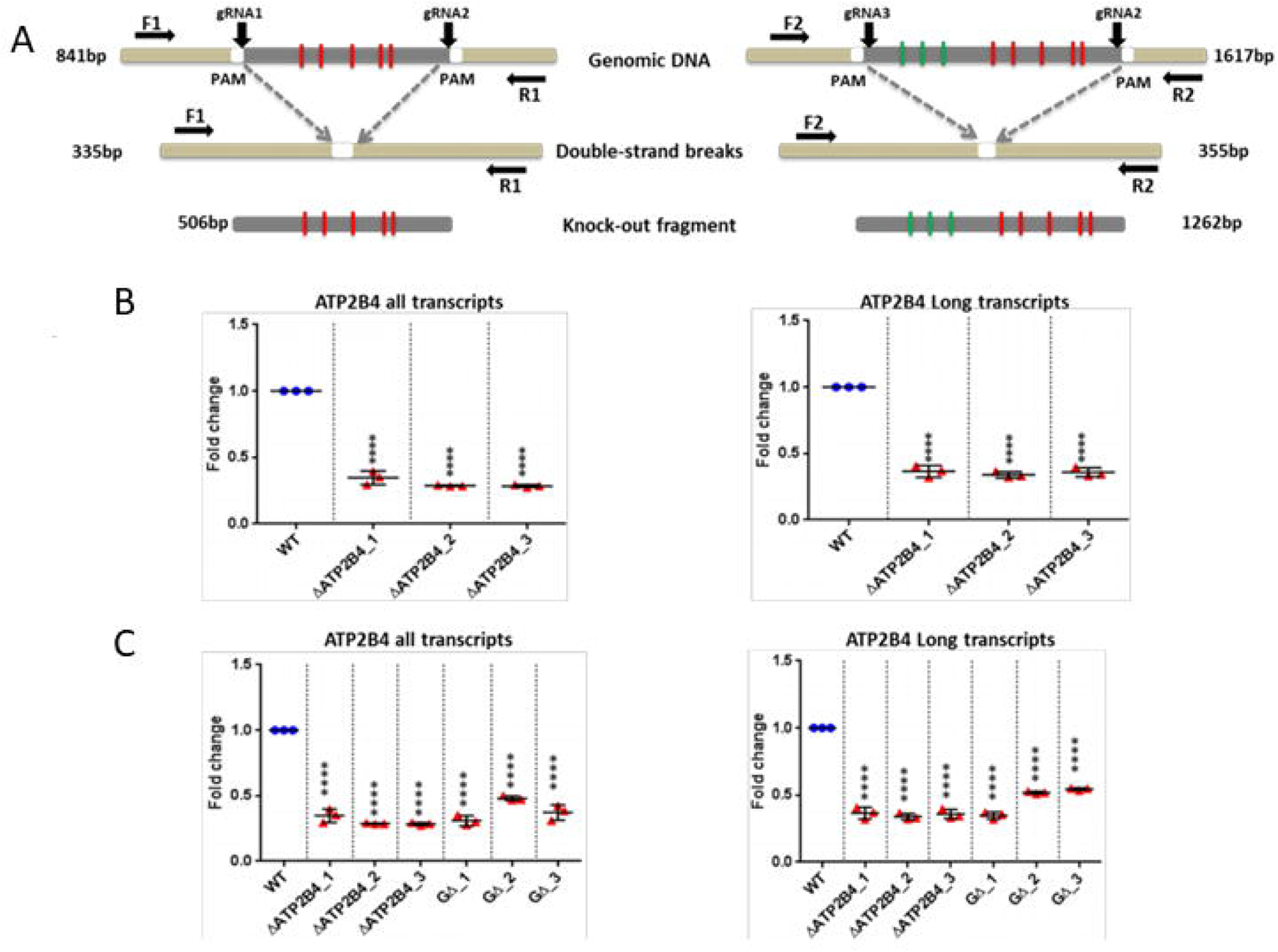
CRISPR-Cas9 mediated genome editing. **(A)** Principle strategy for the generation of DNA knockouts. gRNAs (gRNA1, gRNA2, and gRNA3) were designed flanking the genomic target to delete the two targeted DNA segments comprising either the 5 (506bp) or the 8 (1262bp) candidate SNPs, creating double-strand breaks (DSBs) at 3 bp upstream of the PAM. The resulting DSB is repaired by the NHEJ pathway. The genomic deletion is detected by PCR using primers (F1, R1) and (F2, R2) providing the amplicons of 335bp and 355bp, respectively. **(B)** qPCR analysis of gene expression in wild type K562 cells and the deleted *ATP2B4* clones (for the 5 candidate SNPs) to quantify either all the transcripts or only the two long transcripts of *ATP2B4*. **(C)** qPCR analysis of gene expression in wild type K562 cells and the deleted *ATP2B4* clones (for the 8 SNPs) to quantify either all the transcripts or only the two long transcripts of *ATP2B4*. Error bars show s.d. (*n*=3 independent RNA/cDNA preparations): \*\*\*\**P*<0.0001.

We then deleted a region of 1262-bp to remove the 8 SNPs (rs10751450, rs10751451, rs10751452, rs11240734, rs1541252, rs1541253, rs1541254, and rs1541255) (Fig. 5A) and selected 3 clones validated by sequencing. Clones with this large 1262-bp deletion had a similar reduction in expression compared to the small 506-bp deletion both for total expression and for long transcripts of *ATP2B4*suggesting that the minimal region of 506 bp including (rs11240734, rs1541252, rs1541253, rs1541254, and rs1541255) is enough to regulate the expression of *ATP2B4* (Fig. 5C).

### K562 deleted clones exhibited higher intracellular calcium concentration

Intracellular calcium concentration was measured in 2 clones ΔATP2B4_2 and ΔATP2B4_3 deleted for the region containing rs11240734, rs1541252, rs1541253, rs1541254, and rs1541255 and a representative experiment is shown for each of the clones (Fig. 6A, B). Six and two experiments were performed with ΔATP2B4_2 and ΔATP2B4_3 clones, respectively showing an increase in calcium concentration in deleted clones. Comparing the mean of the MFI yielded significant differences when including only ΔATP2B4_2 clone results (*P*= 0.016) or including both ΔATP2B4_2 and ΔATP2B4_3 results (*P*= 0.002). In the same way, meta-analyses taking into account MFI, SD, and sample size provided evidence of an increase of calcium concentration when including only the results obtained with ΔATP2B4_2 clone (*P*<0.001) or including both ΔATP2B4_2 and ΔATP2B4_3 results (*P*<0.001). Overall, these results indicate that deleted clones expressed *ATP2B4* at a low level, leading to a decrease of calcium efflux and an intracellular accumulation of calcium.

**Figure 6.**
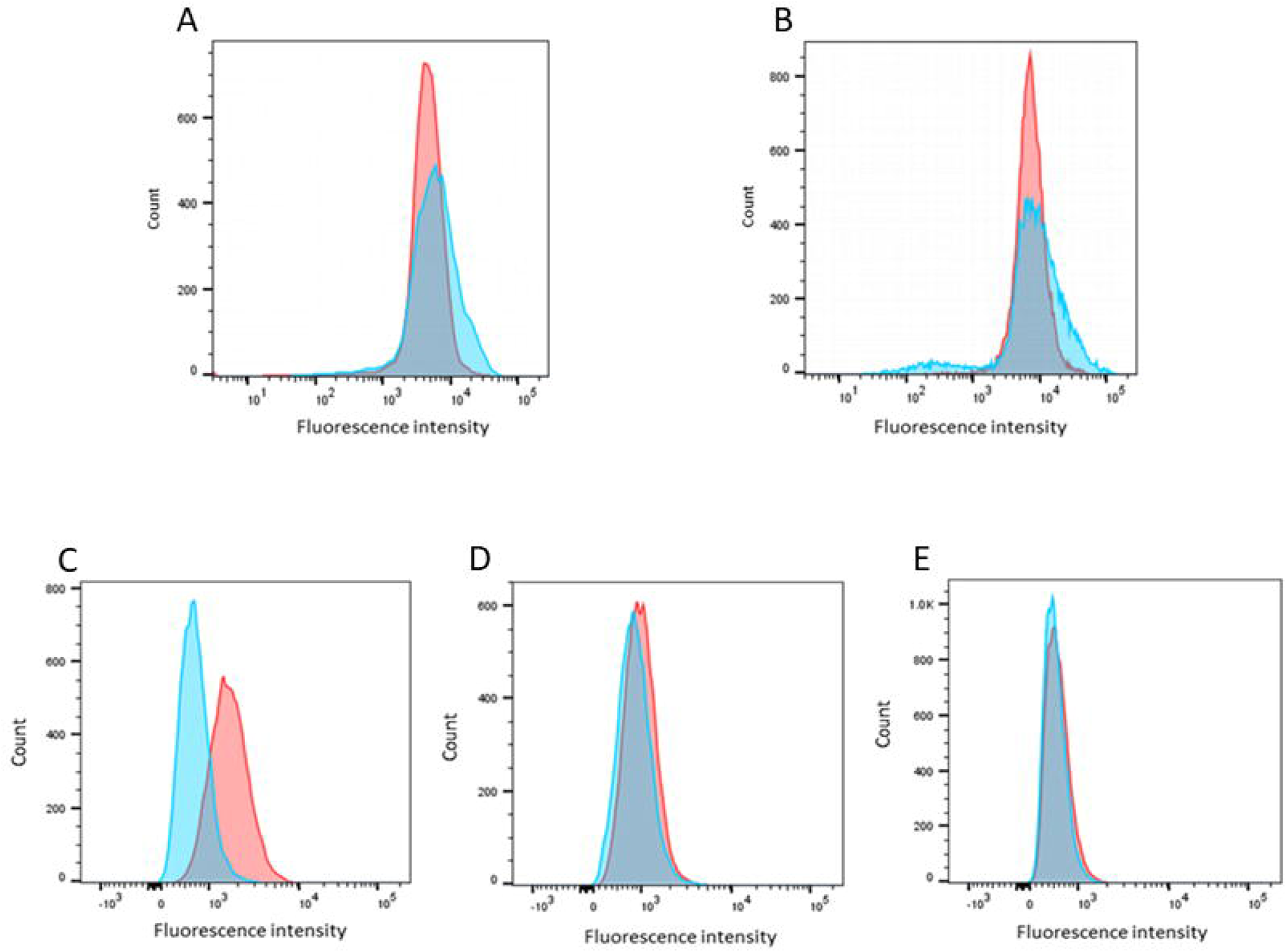
Visualization of calcium concentration and PMCA4 protein expression of K562 cells. **(A, B)** Visualization of calcium concentration of K562 WT (pink) and deleted cells (blue). An increased intracellular calcium concentration was displayed in clone ΔATP2B4_2 **(A)** and, for clone ΔATP2B4_3 **(B)**, respectively. **(C-E)** PMCA4 protein expression using flow cytometry analysis. Representative FACs plot showing isotype (blue) and JA9 (pink) peaks visualized by an APC conjugated secondary antibody in K562 WT cells **(C)**, ΔATP2B4_2 deleted clone **(D)** and ΔATP2B4_3 deleted clone **(E)**. These were visualized using FlowJo software.

### PMCA4 protein is not expressed in K562 deleted clones

The expression of the protein PMCA4 has been assessed in the two deleted clones. Three independent experiments have been performed showing that the PMCA4 protein is not detected in the deleted clones (Fig. 6B, C), whereas this protein is expressed in K562 WT cells (Fig. 6A); MFI difference between WT and deleted clones was significant (*P*=0.002). Meta-analyses that took into account MFI, SD, and sample size further provided evidence of a significant decrease of protein level in deleted clones (*P*<0.0001).

## Discussion

Several GWAS have identified and replicated tagSNPs associated with SM. Most of the causal variants remain, however, unknown. Here we identified 5 *ATP2B4* regulatory SNPs associated with SM in a Senegalese population and in LD with the tagSNPs identified by GWAS (rs4951377 and rs10900585). We showed that these 5 SNPs are located in a promoter region that displays enhancer activity (Epromoter) ^32,33^.

Bioinformatic prioritization of the 125 non-coding SNPs in LD with the tagSNPs associated with SM indicated that the top 10 ranked SNPs by the IW-scoring were on chromosome 1 within *ATP2B4*. When combining the IW-scoring with the annotation of ChIP-seq peaks, rs1541252, rs1541253, rs1541254, and rs1541255 appeared to be the most promising candidates. Interestingly, these SNPs were close to rs11240734 ranked first and fifth based on the IW-score and on the number of ChIP-seq peaks respectively, suggesting that they may be located within a regulatory element. This hypothesis is also supported by 1) the co-location of the SNPs with epigenomic marks associated with promoter or enhancer activity, 2) the characterization of rs11240734, rs1541252, rs1541253, and rs1541254 as eQTLs in blood cells ^34^, 3) the association of rs1541252 and rs1541253 with PMCA4 levels ^35^ and 4) the identification of rs1541255 as regulatory variant based on a reporter assay ^27^.

Consistently, the tagSNPs rs4951377 and rs10900585 ranked 51^st^ and 61^st^ using the IW score, were neither located in a region with histone marks nor with chip-seq peaks suggesting that they are not the causal SNPs. Nevertheless, rs4951377 was an eQTL in blood cells ^34^, illustrating that an eQTL is not necessarily a regulatory variant. The association signal of rs4951377 with gene expression is likely explained by the effect of functional SNPs in LD with it. Besides, we confirmed that rs10751450, rs10751451, rs10751452 ranked 3^rd^, 12^th^, and 13^th^, respectively, could be regulatory SNPs as previously suggested ^23^ and were located within an enhancer altering *ATP2B4* that codes for PMCA4 (Plasma membrane calcium transporting ATPase4) ^23^. This enhancer region is roughly 700 bp away from the region containing rs11240734, rs1541252, rs1541253, rs1541254, and rs1541255, which is potentially a regulatory region according to our results.

Then, we confirmed the association between the tagSNP rs10900585 and SM in a Senegalese population with the same risk genotype and validate its association by meta-analysis integrating all the SM populations ^18,30,31^. In addition, we provided evidence of an association of the 8 SNPs with SM in Senegal. Interestingly, these SNPs except rs10751452 were imputed in the most recent GWAS of SM ^16^ showed a high Bayes Factor value (BF>1.8 107) that was consistent with an association. In the same way, rs1541255 was associated with SM in Kenya ^31^, whereas the other SNPs were not genotyped in this population.

Furthermore, we found a strong haplotype association with SM and we observed that the majority of the Senegalese individuals exhibited either the haplotype combination of all the major alleles or the haplotype combination of all the minor alleles whereas only a few individuals carried other haplotypes. In addition, we showed that this region had a promoter and an enhancer activity and that these activities were modulated by the SNPs and their combination. Interestingly, we observed higher enhancer activity with the haplotype combining the major alleles whereas this haplotype showed lower promoter activity compared to the haplotype with the minor alleles. This result supports a hypothetic model of regulation proposed recently by ^36–38^, whereby genetic variants can impose a regulatory switch between the enhancer and the promoter activity. Our results further suggest that there was an effect of the combination of several SNP alleles on gene expression, thus reflecting the interaction between these SNPs. Although little is known about haplotype’s influence on gene expression, Ying et al ^36^ recently proposed a method to detect eQTL haplotypes and suggested that they are enriched in regulatory regions such as promoters or enhancers. Our finding is also supported by a study providing evidence at numerous loci that “multiple enhancer variants” cooperatively contribute to altered expression of their gene targets and that target transcript levels tend to be modest ^39^. This raised the question of the natural combination of alleles in African populations under selective pressure and supports our finding that the combination of these 5 SNPs was responsible for susceptibility to SM and that there is not a single causal variant.

Also, we showed that the deletion of this regulatory region containing rs11240734, rs1541252, rs1541253, rs1541254, and rs1541255 decreased both the global *ATP2B4* expression and specifically the expression of the long transcripts in the K562 cell line suggesting that it is a promoter with enhancer function (Epromoter). Consistently, decreasing expression of the transcripts in the deleted clones results in the absence of the PMCA4 protein and increased intracellular calcium.

After deleting a region containing rs10751450, rs10751451, and rs10751452, Lessard et al showed a reduction in *ATP2B4* expression and a cytoplasmic calcium accumulation ^23^. Hence, we included these SNPs in our study, and we obtained similar association results when adding rs10751450, rs10751451, and rs10751452 in the haplotype analysis but also a similar decrease in *ATP2B4* expression for the clones either with 5 or 8 SNPs deletion. This finding supports that rs11240734, rs1541252, rs1541253, rs1541254, and rs1541255 affect the *ATP2B4* gene expression as much as rs10751450, rs10751451, and rs10751452 and contribute to development of SM. Here, we have demonstrated the potential impact of these variants in regulating *ATP2B4* expression and intracellular calcium levels. We hypothesize that changes in calcium homeostasis may affect the growth of the parasite in red blood cells and may also affect the clumping of infected red blood cells and their sequestration to the brain microvessels, thus contributing to SM susceptibility. In addition, our results support the hypothesis that a calcium-activated potassium channel may be activated resulting in potassium efflux, dehydration, red blood cell volume loss, increased mean corpuscular hemoglobin concentration, and reduced parasitemia in malaria patients ^23^. Indeed rs1541252 and rs1541255 were found to be associated with mean corpuscular hemoglobin concentration in African-American children and parasitemia in malaria patients, respectively ^31,40^.

Noticeably, it is not excluded that the modulation of intracellular Ca^2+^ concentration alters the physiology of other cell types. For example, the regulation of intracellular Ca^2+^ signaling is a major determinant of CD8+ T cell responsiveness which may have an important role in determining SM. Moreover, the PMCA4 protein is expressed both in the red blood cell and the brain with PMCA4b and PMCA4a the most abundant isoforms in red blood cells and in brain, respectively ^41^. Because these isoforms are tightly and specifically regulated in tissues and cells, we can therefore propose that dysregulation of calcium homeostasis in the brain could also be directly involved in cerebral malaria susceptibility as for different brain disorders such as Alzheimer’s disease and Parkinson’s disease. PMCA4a which is the major isoform present on endothelial cells may also play a crucial role in SM through its role as a negative regulator of vascular endothelial growth factor (VEGF)-activated angiogenesis. In cerebral malaria, the binding of parasitized erythrocytes to the cerebral endothelium and the consequent angiogenic dysregulation play a key role in pathogenesis ^42^. VEGF, a regulator of endothelial inflammation and integrity, is involved in modulating tissue pathology in response to *P. berghei* infection. Indeed, a potent inhibitor to the VEGF signaling pathway aggravated dramatically the course of *P. berghei* infection ^43^.

In conclusion, we identified a new regulatory region that controls the expression of both *ATP2B4* mRNA and PMCA4 protein and that affects intracellular calcium levels. We also showed that the activity of this regulatory region was perturbed by 5 genetic variants associated with SM. This suggests that we identified causal variants within a locus identified through GWAS of SM. This also fosters the development of therapeutic strategies based on the modulation of *ATP2B4* expression and calcium levels. It should be stressed, however, that the effect size of *ATP2B4* was ^4,16,31^. More modest in our study population consistently with previous heritability studies generally, genetic variants in *ATP2B4*, *FREM3-GYP A/B*, *EPHA7*, *ABO*, and *HBB*, which have been identified by several GWAS studies, explain 11% of the genetic contribution to variation in SM susceptibility ^16^. In this way, a polygenic effect has been proposed by Damena et al (2020), suggesting that many genetic variants with a small effect size remain to be discovered ^4^.

## Author contributions

SM and PR designed and supervised the project. SN genotyped most of the genetic variants, whereas MT, AT and FG participated in genotyping. AT extracted the DNA samples in collaboration with RND and performed whole genome amplification. OK, AT and BM were involved in the recruitment, the follow-up of Senegalese individuals, and the collection and interpretation of biological data including parasite density, hematology, and other characteristics from different areas (Dakar and Tambacounda), under the supervision of AD. MT guided CRISPR/Cas9 technology and gene reporter experiments. SN performed qPCR, CRISPR/Cas9, and gene reporter experiments. LB has participated in gene reporter experiments. BP performed cytometry experiments for intracellular calcium levels and membrane protein levels. SN, FR, and PR performed bioinformatic analyses. SN, PR, and SM performed statistical analyses. SS provided useful comments on biological interpretation, whereas SN, SM, and PR interpreted the results and wrote the paper. SM and PR supervised the whole research and provided guidance. All authors have received the manuscript and approved the final version before submitting the article.

## Conflict of Interests

Authors declare no conflict of interests.

## Funding

This work was supported by the African Higher Education Centers of Excellence project (CEA-SAMEF) at UCAD, the Pasteur Institute in Dakar, the Pasteur Institute in Paris, the French embassy in Senegal, INSERM, and Aix-Marseille University. FR and SN were supported by a PhD fellowship from the French ministry of research and the Higher Education Commission (HEC) from Pakistan, respectively.

## Data Availability

All data produced in the present study are available upon reasonable request to the authors

## Abbreviations

ABO: Histo-blood group glycosyltransferases
ATP2B4: ATPase plasma membrane Ca^2+^ transporting 4
eQTL: Expression quantitative trait loci
EPHA7: Ephrin receptor A7
FAC: Fluorescence activated cell sorting
FREM3: FRAS1 related extracellular matrix 3
GYP A/B: Glycophorin A/B
GWAS: Genome-wide association studies
HBB: Hemoglobin subunit beta
IW-scoring: Integrative weighted scoring
LD: Linkage disequilibrium
MFI: Mean fluorescence intensity
OR: Odd ratio
PMCA4: Plasma membrane calcium transporting ATPase4
UCSC: University of California Santa Cruz
VEGF: vascular endothelial growth factor
WT: Wild-type

## Acknowledgments

We thank all the individuals who participated in the study. We thank Iris Manosalva and Charbel Souaid for helpful discussions, and Richard Redon for critical reading of the manuscript. We also thank Sabrina Baaklini and Mai Le Phuong Nguyen for the technical assistance. We thank Dr. Mouhamedou Mansour Fall for his technical assistance and follow-up of patients. We thank Laurence Borge for assistance and the use of the cell culture platform facility (CRCM U1068, Marseille).

## Ethics approval and consent to participate

The Comité d’Ethique de la Recherche de l’Université Cheikh Anta Diop de Dakar has approved the protocols. Each participant or their parents or legal guardians for any minors before inclusion has received written or verbal information in their native language and has given written consent.

## Availability of data and materials

The datasets used and/or analyzed during the current study are available from the corresponding author on reasonable request.

## Appendix A. Supplementary Tables

**Table S1.** Primers sequences for Site-directed mutagenesis of candidate variants for luciferase assay. **Table S2.** Guide RNA and primers sequences for *ATP2B4* knockout via CRISPR-Cas9 genome editing and screening. **Table S3.** Primer sequences (F3/R3) specific for quantification of long transcripts and (F4/R4) for the quantification of all transcripts of *ATP2B4*. **Table S4.** The number of ChIP-seq peaks and IW-scoring rank for each SNP. **Table S5.** Results of the Hardy-Weinberg Equilibrium for *ATP2B4* variants. **Table S6.** Association of *ATP2B4* SNPs with Severe Malaria in the Senegalese population taking age as covariate. **Table S7.** Number of cases and control individuals for each study population used for meta-analysis.

## Appendix B. Supplementary Figure

**Figure S1**. Epigenomic marks in erythroblast.

---

**Figure S1.** Epigenomic marks in erythroblast. Visual representation of the position of 5 *ATP2B4* candidate variants (rs11240734, rs1541252, rs1541253, rs1541254, and rs1541255; encircled red) and 3 previously identified regulatory variants (rs10751450, rs10751451 and rs10751452; encircled blue) on chromosome 1, located within a DNaseI hypersensitivity region and peaks of H3K4me3, H3K4me1 and H3K27ac histone marks in erythroblast.

## Notes

### Competing Interest Statement

The authors have declared no competing interest.

### Author Declarations

Ethics committee of the Universite Cheikh Anta Diop (UCAD)

## References

1. Mackinnon MJ, Mwangi TW, Snow RW, Marsh K, Williams TN. Heritability of malaria in Africa. PLoS Med. 2005;2(12):e340.

2. Miller LH, Baruch DI, Marsh K, Doumbo OK. The pathogenic basis of malaria. Nature. 2002;415(6872):673–679.

3. Weatherall D, Clegg J. Genetic variability in response to infection: malaria and after. Genes & Immunity. 2002;3(6):331–337.

4. Damena D, Chimusa ER. Genome-wide heritability analysis of severe malaria resistance reveals evidence of polygenic inheritance. Human molecular genetics. 2020;29(1):168–176.

5. Rihet P, Traoré Y, Abel L, Aucan C, Traoré-Leroux T, Fumoux F. Malaria in humans: Plasmodium falciparum blood infection levels are linked to chromosome 5q31-q33. The American Journal of Human Genetics. 1998;63(2):498–505.

6. Brisebarre A, Kumulungui B, Sawadogo S, et al. A genome scan for Plasmodium falciparum malaria identifies quantitative trait loci on chromosomes 5q31, 6p21. 3, 17p12, and 19p13. Malaria journal. 2014;13(1):1–7.

7. Flori L, Sawadogo S, Esnault C, Delahaye NF, Fumoux F, Rihet P. Linkage of mild malaria to the major histocompatibility complex in families living in Burkina Faso. Hum Mol Genet. 2003;12(4):375–378.

8. Jepson A, Sisay-Joof F, Banya W, et al. Genetic linkage of mild malaria to the major histocompatibility complex in Gambian children: study of affected sibling pairs. Bmj. 1997;315(7100):96–97.

9. Flori L, Kumulungui B, Aucan C, et al. Linkage and association between Plasmodium falciparum blood infection levels and chromosome 5q31–q33. Genes & Immunity. 2003;4(4):265–268.

10. Garcia A, Marquet S, Bucheton B, et al. Linkage analysis of blood Plasmodium falciparum levels: interest of the 5q31-q33 chromosome region. The American journal of tropical medicine and hygiene. 1998;58(6):705–709.

11. Milet J, Nuel G, Watier L, et al. Genome wide linkage study, using a 250K SNP map, of Plasmodium falciparum infection and mild malaria attack in a Senegalese population. PLoS One. 2010;5(7):e11616.

12. Sakuntabhai A, Ndiaye R, Casadémont I, et al. Genetic determination and linkage mapping of Plasmodium falciparum malaria related traits in Senegal. PLoS One. 2008;3(4):e2000.

13. Band G, Le QS, Jostins L, et al. Imputation-based meta-analysis of severe malaria in three African populations. PLoS Genet. 2013;9(5):e1003509.

14. Jallow M, Teo YY, Small KS, et al. Genome-wide and fine-resolution association analysis of malaria in West Africa. Nature genetics. 2009;41(6):657–665.

15. Network MGE. A novel locus of resistance to severe malaria in a region of ancient balancing selection. Nature. 2015;526(7572):253–257.

16. Network MGE. Insights into malaria susceptibility using genome-wide data on 17,000 individuals from Africa, Asia and Oceania. Nature communications. 2019;10.

17. Ravenhall M, Campino S, Sepúlveda N, et al. Novel genetic polymorphisms associated with severe malaria and under selective pressure in North-eastern Tanzania. PLoS genetics. 2018;14(1):e1007172.

18. Timmann C, Thye T, Vens M, et al. Genome-wide association study indicates two novel resistance loci for severe malaria. Nature. 2012;489(7416):443–446.

19. Pule G, Chimusa E, Mnika K, et al. Beta-globin gene haplotypes and selected Malaria-associated variants among black Southern African populations. Global health, epidemiology and genomics. 2017;2.

20. Leffler EM, Band G, Busby GB, et al. Resistance to malaria through structural variation of red blood cell invasion receptors. Science. 2017;356(6343).

21. Mayer DG, Cofie J, Jiang L, et al. Glycophorin B is the erythrocyte receptor of Plasmodium falciparum erythrocyte-binding ligand, EBL-1. Proceedings of the National Academy of Sciences. 2009;106(13):5348–5352.

22. Sim B, Chitnis C, Wasniowska K, Hadley T, Miller L. Receptor and ligand domains for invasion of erythrocytes by Plasmodium falciparum. Science. 1994;264(5167):1941–1944.

23. Lessard S, Gatof ES, Beaudoin M, et al. An erythroid-specific ATP2B4 enhancer mediates red blood cell hydration and malaria susceptibility. The Journal of clinical investigation. 2017;127(8):3065–3074.

24. Thiam A, Baaklini S, Mbengue B, et al. NCR3 polymorphism, haematological parameters, and severe malaria in Senegalese patients. PeerJ. 2018;6:e6048.

25. Wang J, Dayem Ullah AZ, Chelala C. IW-Scoring: an Integrative Weighted Scoring framework for annotating and prioritizing genetic variations in the noncoding genome. Nucleic acids research. 2018;46(8):e47–e47.

26. Chèneby J, Gheorghe M, Artufel M, Mathelier A, Ballester B. ReMap 2018: an updated atlas of regulatory regions from an integrative analysis of DNA-binding ChIP-seq experiments. Nucleic acids research. 2018;46(D1):D267–D275.

27. van Arensbergen J, Pagie L, FitzPatrick VD, et al. High-throughput identification of human SNPs affecting regulatory element activity. Nature genetics. 2019;51(7):1160.

28. Martorell-Marugan J, Toro-Dominguez D, Alarcon-Riquelme ME, Carmona-Saez P. MetaGenyo: a web tool for meta-analysis of genetic association studies. BMC bioinformatics. 2017;18(1):563.

29. Wallace BC, Dahabreh IJ, Trikalinos TA, Lau J, Trow P, Schmid CH. Closing the gap between methodologists and end-users: R as a computational back-end. J Stat Softw. 2012;49(5):1–15.

30. Malaria Genomic Epidemiology N, Malaria Genomic Epidemiology N. Reappraisal of known malaria resistance loci in a large multicenter study. Nat Genet. 2014;46(11):1197–1204.

31. Ndila CM, Uyoga S, Macharia AW, et al. Human candidate gene polymorphisms and risk of severe malaria in children in Kilifi, Kenya: a case-control association study. Lancet Haematol. 2018;5(8):e333–e345.

32. Dao LT, Galindo-Albarrán AO, Castro-Mondragon JA, et al. Genome-wide characterization of mammalian promoters with distal enhancer functions. Nature genetics. 2017;49(7):1073.

33. Dao LT, Spicuglia S. Transcriptional regulation by promoters with enhancer function. Transcription. 2018;9(5):307–314.

34. Westra H-J, Peters MJ, Esko T, et al. Systematic identification of trans eQTLs as putative drivers of known disease associations. Nature genetics. 2013;45(10):1238–1243.

35. Zámbó B, Várady G, Padányi R, et al. Decreased calcium pump expression in human erythrocytes is connected to a minor haplotype in the ATP2B4 gene. Cell Calcium. 2017;65:73–79.

36. Gao P, Xia J-H, Sipeky C, et al. Biology and clinical implications of the 19q13 aggressive prostate cancer susceptibility locus. Cell. 2018;174(3):576–589. e518.

37. Hua JT, Ahmed M, Guo H, et al. Risk SNP-mediated promoter-enhancer switching drives prostate cancer through lncRNA PCAT19. Cell. 2018;174(3):564–575. e518.

38. Ying D, Li MJ, Sham PC, Li M. A powerful approach reveals numerous expression quantitative trait haplotypes in multiple tissues. Bioinformatics. 2018;34(18):3145–3150.

39. Corradin O, Saiakhova A, Akhtar-Zaidi B, et al. Combinatorial effects of multiple enhancer variants in linkage disequilibrium dictate levels of gene expression to confer susceptibility to common traits. Genome research. 2014;24(1):1–13.

40. Li J, Glessner JT, Zhang H, et al. GWAS of blood cell traits identifies novel associated loci and epistatic interactions in Caucasian and African-American children. Human molecular genetics. 2013;22(7):1457–1464.

41. Caride AJ, Filoteo AG, Penniston JT, Strehler EE. The Plasma Membrane Ca2+ Pump Isoform 4a Differs from Isoform 4b in the Mechanism of Calmodulin Binding and Activation Kinetics IMPLICATIONS FOR Ca2+ SIGNALING. Journal of Biological Chemistry. 2007;282(35):25640–25648.

42. Furuta T, Kimura M, Watanabe N. Elevated levels of vascular endothelial growth factor (VEGF) and soluble vascular endothelial growth factor receptor (VEGFR)-2 in human malaria. The American journal of tropical medicine and hygiene. 2010;82(1):136–139.

43. Canavese M, Crisanti A. Vascular endothelial growth factor (VEGF) and lovastatin suppress the inflammatory response to Plasmodium berghei infection and protect against experimental cerebral malaria. Pathogens and global health. 2015;109(6):266–274.

