## Supplementary Tables for "Identification of *ATP2B4* regulatory element containing functional genetic variants associated with severe malaria"

**Table S1:** Primers sequences for Site-directed mutagenesis of candidate variants for luciferase assay.

| **SNP** | **Forward Primer** | **Reverse Primer** |
| --- | --- | --- |
| rs11240734 (T/C) | ATACTTATGGTTGCTCTAATGGTTTCTTTTTG | CCAGCATCTGCAAGGGCA |
| rs1541252 (C/T) | TCAGGCCTAGCTATCAGTTCAG | CGAGTAGCCGTCCGAAGTC |
| rs1541253 (C/T) | CCTGGACAACCACTATCATCAC | GTAGGTAGGTTCCTAGCTTAG |
| rs1541254 (G/C) | AGACTTCATAGGGAAGAAAGGATCTAGAC | GGCAGCGAGAGGAGGAAG |
| rs1541255 (A/G) | GACTTCATACAGAAGAAAGGATCTAGACTTCG | TGGCAGCGAGAGGAGGAA |

**Table S2:** Guide RNA and primers sequences for *ATP2B4* knockout via CRISPR-Cas9 genome editing and screening.

| **Guide RNA** | **Sequence 5’-3’** | **Coordinates (hg19)** |
| --- | --- | --- |
| gRNA1 | TCCTCTACATTGGAGTTTAC AGG | chr1:203651648-203651667 |
| gRNA2 | TAGACTTCGGACGGCTACTC GGG | chr1:203652154-203652173 |
| gRNA3 | AGACTCTGGTGCTCACTCAC AGG | chr1:203650890-203650909 |
| F1 | GGCCACCCTTCAGATCACTT | chr1:203651422-203651441 |
| R1 | GCCTCCCTGTCTCAACTTCT | chr1:203652243-203652262 |
| F2 | CTGAAACTGAGAAGGGGCCT | chr1:203650729-203650748 |
| R2 | TGGGTTCGTCATTTTGCCTG | chr1:203652326-203652345 |

**Table S3:** Primer sequences (F3/R3) specific for quantification of long transcripts and (F4/R4) for the quantification of all transcripts of *ATP2B4*.

| **Guide RNA** | **Sequence 5’-3’** | **Coordinates (hg19)** |
| --- | --- | --- |
| ATP2B4_del_F3 | CAAGAGTCTGGCCCGAGTTA | chr1:203595946-203595965 |
| ATP2B4_del_R3 | TCTGCTGTTGAGATCGTCCA | chr1:203596006-203596025 |
| ATP2B4_del_F4 | TTGGAGAACTTCTGTGGGGC | chr1:203693094-203693113 |
| ATP2B4_del_R4 | AGGAGATCACCAAGGATGCC | chr1:203696590-203696609 |

**Table S4:** Number of ChIP-seq peaks and IW-scoring rank for each SNP

| **rsID** | **Genomic coordinates** | **IW-score** | **IW scoring Rank** | **Number of ChIP-seq peaks** | **eQTL hits** |
| --- | --- | --- | --- | --- | --- |
| rs11240734 | chr1: 203682696 | 3,4634 | 1 | 54 | 3 hits |
| rs1541252 | chr1: 203682799 | 3,1663 | 2 | 90 | 3 hits |
| rs10751450 | chr1: 203681817 | 3,1011 | 3 | 46 |  |
| rs2228445 | chr1: 203698281 | 2,7194 | 4 | 18 | 3 hits |
| rs202111522 | chr1: 203682602 | 2,4339 | 5 | 43 |  |
| rs1541254 | chr1: 203683012 | 2,5458 | 6 | 104 |  |
| rs1541255 | chr1: 203683013 | 1,8505 | 7 | 103 |  |
| rs10594838 | chr1: 203685540 | 1,7405 | 8 | 2 |  |
| rs10625220 | chr1: 203686622 | -0,041 | 9 | 15 |  |
| rs1541253 | chr1: 203682912 | 1,1953 | 10 | 149 |  |
| rs8176719 | chr9: 133257521 | 2,482 | 11 | 7 | 14 hits |
| rs10751451 | chr1: 203681850 | 1,0658 | 12 | 49 | 3 hits |
| rs10751452 | chr1: 203681902 | 0,7364 | 13 | 50 |  |
| rs1999063 | chr6: 92558037 | -0,1021 | 14 | 0 |  |
| rs200499024 | chr1: 203681934 | 0,0246 | 15 | 50 |  |
| rs11240733 | chr1: 203681464 | -0,2697 | 16 | 13 |  |
| rs6594007 | chr1: 203685644 | -0,3419 | 17 | 2 | 3 hits |
| rs184527160 | chr4: 143782987 | -0,3847 | 18 | 8 |  |
| rs4951369 | chr1: 203683555 | -0,4386 | 19 | 0 |  |
| rs192832154 | chr4: 143712082 | -0,3659 | 20 | 10 |  |
| rs10736845 | chr1: 203681658 | -0,4748 | 21 | 27 | 3 hits |
| rs4951070 | chr1: 203683570 | -0,5606 | 22 | 0 | 3 hits |
| rs1419114 | chr1: 203683316 | -1,218 | 23 | 16 | 3 hits |
| rs115426616 | chr4: 143829455 | -0,7402 | 24 | 5 |  |
| rs10751449 | chr1: 203681656 | -0,852 | 25 | 27 |  |
| rs7551442 | chr1: 203685993 | -1,098 | 26 | 5 | 3 hits |
| rs11240731 | chr1: 203681208 | -0,9579 | 27 | 6 |  |
| rs145677206 | chr4: 143714067 | -1,1407 | 28 | 6 |  |
| rs7551560 | chr1: 203686142 | -1,1388 | 29 | 13 | 3 hits |
| rs184063934 | chr4: 143712636 | -1,2999 | 30 | 2 |  |
| rs76471583 | chr4: 143682249 | -1,3356 | 31 | 0 |  |
| rs4951381 | chr1: 203691710 | -1,462 | 32 | 13 | 3 hits |
| rs4951074 | chr1: 203691653 | -1,39 | 33 | 14 | 3 hits |
| rs7539122 | chr1: 203685915 | -1,4746 | 34 | 2 | 3 hits |
| rs183974736 | chr4: 143820703 | -1,5281 | 35 | 3 |  |
| rs141787228 | chr6: 92561006 | -1,5312 | 36 | 0 |  |
| rs4951081 | chr1: 203702749 | -1,7027 | 37 | 3 | 2 hits |
| rs146104413 | chr4: 143727261 | -1,6695 | 38 | 0 |  |
| rs7546390 | chr1: 203680295 | -1,8079 | 39 | 0 |  |
| rs6692632 | chr1: 203685418 | -1,7525 | 40 | 3 |  |
| rs143128154 | chr1: 203687259 | -1,2236 | 41 | 3 |  |
| rs181423417 | chr4: 143757218 | -1,9533 | 42 | 0 |  |
| rs201385421 | chr1: 203680619 | -2,1083 | 43 | 1 |  |
| rs186873296 | chr4: 143781321 | -2,1521 | 44 | 4 |  |
| rs6594006 | chr1: 203685610 | -2,1009 | 45 | 2 | 3 hits |
| rs6658130 | chr1: 203685440 | -2,2154 | 46 | 3 | 3 hits |
| rs182386242 | chr4: 143781808 | -2,2226 | 47 | 6 |  |
| rs10900588 | chr1: 203687686 | -2,1508 | 48 | 1 | 3 hits |
| rs142731296 | chr4: 143721083 | -2,3307 | 49 | 0 |  |
| rs148995556 | chr4: 143834361 | -2,3027 | 50 | 3 |  |
| rs10900585 | chr1: 203684896 | -2,392 | 51 | 0 |  |
| rs184820425 | chr4: 143703360 | -2,3098 | 52 | 0 |  |
| rs189887464 | chr4: 143814353 | -2,343 | 53 | 0 |  |
| rs189149534 | chr4: 143735453 | -2,368 | 54 | 3 |  |
| rs190618313 | chr4: 143683056 | -2,3782 | 55 | 2 |  |
| rs2365859 | chr1: 203688560 | -2,3568 | 56 | 6 |  |
| rs149914432 | chr4: 143745525 | -2,4268 | 57 | 0 |  |
| rs6697384 | chr1: 203708437 | -2,4504 | 58 | 1 |  |
| rs12035565 | chr1: 203680750 | -2,3356 | 59 | 0 |  |
| rs4951377 | chr1: 203689343 | -2,4195 | 60 | 0 | 3 hits |
| rs184497846 | chr4: 143826118 | -2,4907 | 61 | 0 |  |
| rs4951378 | chr1: 203689654 | -2,5181 | 62 | 0 | 3 hits |
| rs4951375 | chr1: 203686657 | -2,689 | 63 | 7 | 3 hits |
| rs4707740 | chr6: 92475692 | -2,5897 | 64 | 0 |  |
| rs151041851 | chr4: 143709358 | -2,6522 | 65 | 0 |  |
| rs188245886 | chr4: 143685417 | -2,6877 | 66 | 0 |  |
| rs10900589 | chr1: 203687846 | -2,7977 | 67 | 1 | 3 hits |
| rs2183001 | chr6: 92559279 | -2,755 | 68 | 11 |  |
| rs183414953 | chr4: 143687478 | -2,7972 | 69 | 0 |  |
| rs147037418 | chr4: 143783021 | -2,8153 | 70 | 8 |  |
| rs4707739 | chr6: 92475163 | -2,8479 | 71 | 0 |  |
| rs4951370 | chr1: 203683777 | -2,8554 | 72 | 0 |  |
| rs184627523 | chr4: 143813546 | -2,9012 | 73 | 1 |  |
| rs151007681 | chr4: 143693393 | -2,9308 | 74 | 0 |  |
| rs2365860 | chr1: 203687102 | -2,9416 | 75 | 3 | 3 hits |
| rs6594008 | chr1: 203685723 | -2,8381 | 76 | 0 | 3 hits |
| rs6594011 | chr1: 203686827 | -3,0051 | 77 | 0 |  |
| rs142537619 | chr4: 143740252 | -3,0594 | 78 | 0 |  |
| rs7546599 | chr1: 203680540 | -3,2011 | 79 | 1 |  |
| rs186896080 | chr4: 143707921 | -3,0982 | 80 | 0 |  |
| rs4951371 | chr1: 203683818 | -3,1091 | 81 | 0 |  |
| rs144794297 | chr4: 143681765 | -3,107 | 82 | 0 |  |
| rs10793762 | chr1: 203684970 | -3,0229 | 83 | 0 |  |
| rs7547793 | chr1: 203684416 | -3,1917 | 84 | 0 |  |
| rs4347266 | chr1: 203687050 | -3,0943 | 85 | 0 |  |
| rs187764080 | chr4: 143694377 | -3,1214 | 86 | 0 |  |
| rs186094021 | chr4: 143784433 | -3,1403 | 87 | 0 |  |
| rs148311540 | chr4: 143831996 | -3,1777 | 88 | 0 |  |
| rs146553388 | chr4: 143678652 | -3,2106 | 89 | 0 |  |
| rs184114500 | chr4: 143716593 | -3,2087 | 90 | 0 |  |
| rs12122293 | chr1: 203687989 | -3,2423 | 91 | 0 |  |
| rs192804806 | chr4: 143814875 | -3,271 | 92 | 0 |  |
| rs187377484 | chr4: 143840986 | -3,2947 | 93 | 1 |  |
| rs6696548 | chr1: 203686939 | -3,3281 | 94 | 0 |  |
| rs10900586 | chr1: 203684957 | -3,3152 | 95 | 0 |  |
| rs182977569 | chr4: 143707466 | -3,3377 | 96 | 1 |  |
| rs2365858 | chr1: 203688621 | -3,3986 | 97 | 7 | 3 hits |
| rs3851298 | chr1: 203695919 | -3,3045 | 98 | 0 |  |
| rs147565832 | chr4: 143764458 | -3,3684 | 99 | 0 |  |
| rs116706045 | chr4: 143828598 | -3,349 | 100 | 0 |  |
| rs7555703 | chr1: 203684562 | -3,325 | 101 | 0 |  |
| rs6594010 | chr1: 203686754 | -3,3861 | 102 | 0 |  |
| rs145945511 | chr4: 143785347 | -3,3956 | 103 | 0 |  |
| rs143493147 | chr4: 143823880 | -3,3901 | 104 | 0 |  |
| rs141987247 | chr4: 143687266 | -3,4094 | 105 | 0 |  |
| rs138428794 | chr4: 143732434 | -3,4284 | 106 | 0 |  |
| rs7514742 | chr1: 203680588 | -3,5388 | 107 | 1 |  |
| rs149060195 | chr4: 143732408 | -3,5054 | 108 | 0 |  |
| rs9345242 | chr6: 92517195 | -3,519 | 109 | 0 |  |
| rs6692627 | chr1: 203688092 | -3,5304 | 110 | 0 |  |
| rs72926662 | chr6: 92598572 | -3,5808 | 111 | 0 |  |
| rs184908374 | chr4: 143744600 | -3,5872 | 112 | 0 |  |
| rs186790584 | chr4: 143758987 | -3,6109 | 113 | 1 |  |
| rs183166203 | chr4: 143734505 | -3,6343 | 114 | 0 |  |
| rs62418762 | chr6: 92508980 | -3,6525 | 115 | 0 |  |
| rs189723550 | chr4: 143764153 | -3,6609 | 116 | 0 |  |
| rs192563008 | chr4: 143824276 | -3,6974 | 117 | 0 |  |
| rs10944597 | chr6: 92494332 | -3,6797 | 118 | 1 |  |
| rs77397300 | chr4: 143752045 | -3,7663 | 119 | 0 |  |
| rs185357240 | chr4: 143779807 | -3,7691 | 120 | 1 |  |
| rs187621111 | chr4: 143687489 | -3,8762 | 121 | 0 |  |
| rs9353916 | chr6: 92480916 | -3,8775 | 122 | 1 |  |
| rs7535676 | chr1: 203684658 | -3,9418 | 123 | 0 |  |
| rs184895969 | chr4: 143777375 | -3,9361 | 124 | 1 |  |
| rs147881070 | chr4: 143747988 | -4,0485 | 125 | 0 |  |

**Table S5:** Results of the Hardy-Weinberg Equilibrium for *ATP2B4* variants.

| **SNP** | **H-W (*P*-value)** |
| --- | --- |
| rs10900585 | 0.52 |
| rs11240734 | 0.27 |
| rs1541252 | 0.27 |
| rs1541253 | 0.27 |
| rs1541254 | 0.77 |
| rs1541255 | 0.27 |
| rs10751450 | 0.76 |
| rs10751451 | 0.53 |
| rs10751152 | 0.76 |

**Table S6:** Association of *ATP2B4* SNPs with Severe Malaria in the Senegalese population. *P*-values and OR was calculated according to logistic regression test taking age as covariate.

| **SNP** | **Position^a^** | **Minor Allele** | **MAF^b^** | **Risk Genotype** | **Controls %** | **Severe Malaria %** | **OR** | **95% CI** | ***P*** |
| --- | --- | --- | --- | --- | --- | --- | --- | --- | --- |
| rs10900585 (T>G) | 203684896 | G | 0.41 | TT | 32.9 | 48.7 | 1.82 | 0.99-3.34 | 0.055 |
| rs11240734 (T>C) | 203682696 | C | 0.40 | TT | 31.6 | 53.8 | 2.38 | 1.29-4.40 | 0.006 |
| rs1541252 (C>T) | 203682799 | T | 0.40 | CC | 31.6 | 53.8 | 2.38 | 1.29-4.40 | 0.006 |
| rs1541253 (C>T) | 203682912 | T | 0.40 | CC | 31.6 | 53.8 | 2.38 | 1.29-4.40 | 0.006 |
| rs1541254 (G>C) | 203683012 | C | 0.43 | GG | 31.6 | 53.8 | 2.38 | 1.29-4.40 | 0.006 |
| rs1541255 (A>G) | 203683013 | G | 0.40 | AA | 31.6 | 53.8 | 2.38 | 1.29-4.40 | 0.006 |
| rs10751450 (C>T) | 203681817 | T | 0.41 | CC | 32.9 | 50.4 | 1.98 | 1.06-3.67 | 0.030 |
| rs10751451 (C>T) | 203681850 | T | 0.40 | CC | 34.2 | 56.4 | 2.23 | 1.21-4.12 | 0.011 |
| rs10751452 (T>C) | 203681902 | C | 0.39 | TT | 35.4 | 55.6 | 2.05 | 1.11-3.78 | 0.021 |

^a^ Data represent the position on chromosome 1 according to human hg38 coordinates

^b^ MAF (Minor allele frequency) was estimated from Senegalese healthy controls.

OR, odds ratio; CI, confidence interval.

**Table S7:** Number of cases and control individuals for each study population used for meta-analysis

| **Reference** | **PMID** | **Year** | **Population** | **No. of cases** | **No. of controls** | **Genotype (cases)** | | | **Genotype (control)** | | |
| --- | --- | --- | --- | --- | --- | --- | --- | --- | --- | --- | --- |
|  |  |  |  |  |  | **TT** | **TG** | **GG** | **TT** | **TG** | **GG** |
| Rockett *et al.* | 25261933 | 2014 | Mali | 427 | 322 | 228 | 164 | 35 | 156 | 141 | 25 |
| Rockett *et al.* | 25261933 | 2014 | Burkina Faso | 843 | 720 | 388 | 377 | 78 | 365 | 292 | 63 |
| Rockett *et al.* | 25261933 | 2014 | Ghana(Navrongo) | 614 | 165 | 334 | 246 | 34 | 86 | 74 | 5 |
| Rockett *et al.* | 25261933 | 2014 | Ghana(Kumasi) | 761 | 1231 | 294 | 365 | 102 | 405 | 586 | 240 |
| Rockett *et al.* | 25261933 | 2014 | Nigeria | 76 | 34 | 25 | 39 | 12 | 15 | 15 | 4 |
| Rockett *et al.* | 25261933 | 2014 | Kenya | 1624 | 3762 | 688 | 786 | 150 | 1644 | 1689 | 429 |
| Rockett *et al.* | 25261933 | 2014 | Tanzania | 425 | 451 | 178 | 199 | 48 | 182 | 211 | 58 |
| Rockett *et al.* | 25261933 | 2014 | Malawi | 1374 | 2562 | 654 | 595 | 125 | 1159 | 1105 | 298 |
| Rockett *et al.* | 25261933 | 2014 | Vietnam | 779 | 2456 | 746 | 33 | 0 | 2367 | 89 | 0 |
| Ndila *et al.* | 30033078 | 2018 | Kenya | 2200 | 3762 | 989 | 973 | 238 | 1639 | 1688 | 435 |
| Timmann *et al.* | 22895189 | 2012 | Ghana (GWA) | 1325 | 828 | 510 | 624 | 191 | 269 | 406 | 153 |
| Timmann *et al.* | 22895189 | 2012 | Ghana (replica) | 1320 | 2222 | 507 | 622 | 191 | 697 | 1095 | 430 |
| Timmann *et al.* | 22895189 | 2012 | The Gambia | 909 | 1304 | 408 | 402 | 99 | 517 | 608 | 179 |
| Nisar *et al.* |  | 2021 | Senegal | 117 | 79 | 57 | 44 | 16 | 26 | 41 | 12 |
