## Supplementary figures and images for "Identification of *ATP2B4* regulatory element containing functional genetic variants associated with severe malaria"

### Supplementary Figure

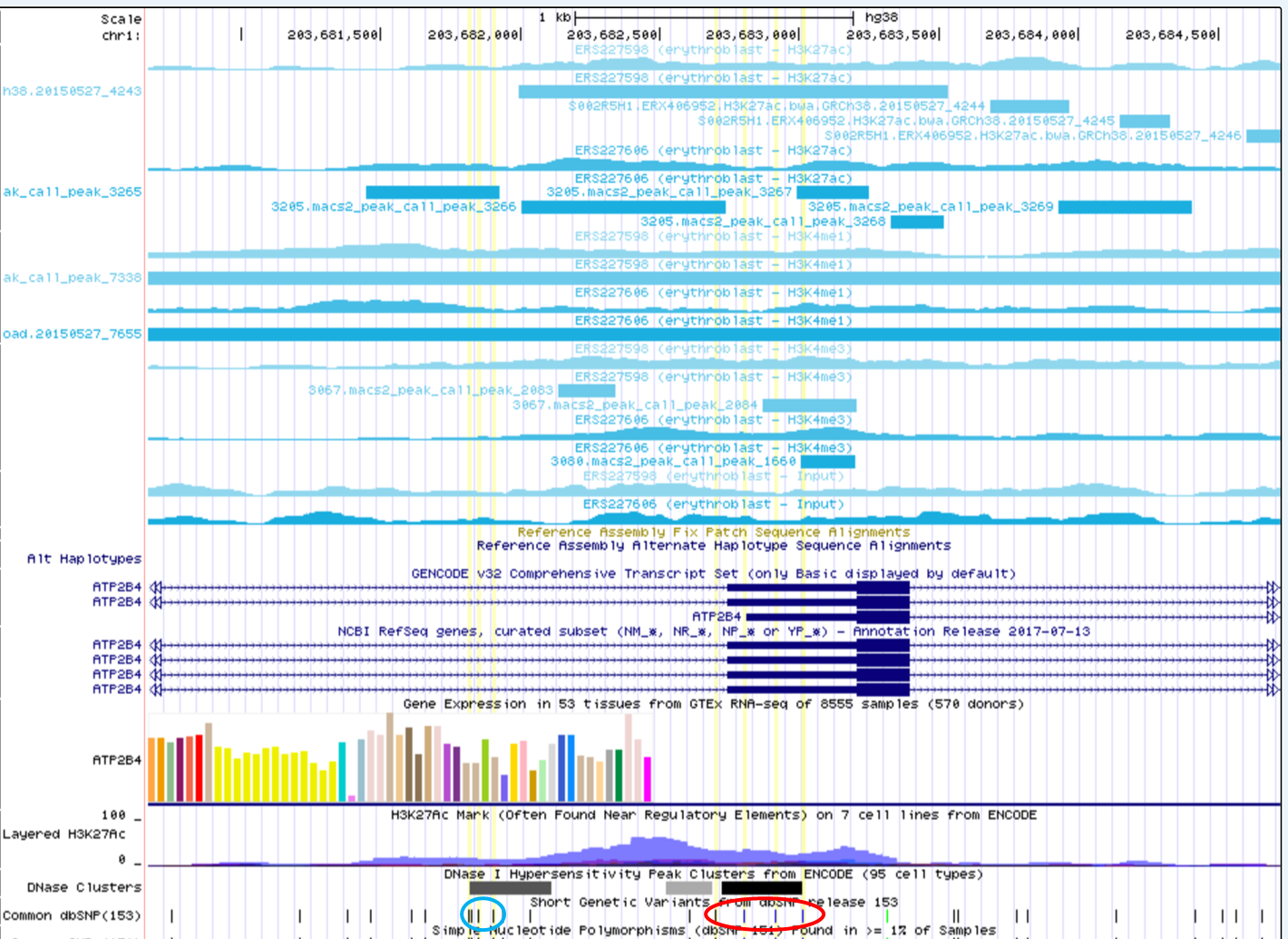
